# Salivary proteome of aphthous stomatitis reveals the participation of vitamin metabolism, nutrients, and bacteria

**DOI:** 10.1101/2021.08.01.21261411

**Authors:** Romina Hernández-Olivos, Mariagrazia Muñoz, Esteban Núñez, Paola Andrea Camargo-Ayala, Jenaro Garcia-Huidobro, Alfredo Pereira, Fabiane M. Nachtigall, Leonardo S. Santos, César Rivera

## Abstract

There are currently no preventative options for recurrent aphthous stomatitis, and the only available treatments are palliative. This is partly due to a poor understanding of its etiopathogenesis. In this case-control study, we characterized the salivary proteome of patients with recurrent aphthous stomatitis in the presence and absence of lesions. Through mass spectrometry-based proteomics and bioinformatics tools, we identified that the presence of oral ulcers is associated with several specific biological processes, including the metabolic pathways of vitamin B9, B12, nitrogen, selenium, and the bacterium *Neisseria meningitidis*. These changes occurred only in the presence of clinically visible lesions, and there were no relevant differences between patients in anatomical regions unaffected by ulcers. Additionally, using western blot and ELISA assays, we verified that carbonic anhydrase 1 (CA1, UniProtKB - P00915, CAH1_HUMAN) and hemoglobin subunit beta (HBB, UniProtKB - P68871, HBB_HUMAN) proteins are highly expressed during the ulcerative and remission phases of recurrent aphthous stomatitis. Our results cumulatively support saliva as an indicator of the pathophysiological changes, which occur during the clinical course of lesions. From a clinical perspective, we suggest that recurrent aphthous stomatitis is a condition triggered by temporary biological changes in people with lesions.

## INTRODUCTION

In oral medicine, there are several conditions which are constituted by recurring episodes of unidentified causation ^1^. Subsequently, available therapies lack specificity, and are of questionable efficacy. The largest representative of this group is recurrent aphthous stomatitis (also known as canker sores or recurrent aphthae), the most common ulcerative condition of the oral mucosa worldwide ^2^. It is characterized by recurrent episodes of painful ulcerations lacking any association with systemic disease ^3^. Owing to its painful nature, this disease can compromise important functions such as nutrition, speech and oral hygiene ^4^, as well as affecting quality of life ^5^. This is a critical health issue, as the ulcers can persist for over 2 weeks, with recurring episodes between 1 and 4 months. ^6^. Although other oral diseases such as periodontal disease and caries are more prevalent, patients with recurrent aphthous stomatitis cannot prevent the development of lesions, and only palliative therapeutic alternatives (corticosteroid preparations) are available. Despite the increasing use of topical corticosteroids for recurrent aphthous stomatitis over the past several years, high-quality evidence of their efficacy is lacking ^7^, and relief is often only symptomatic with no effect on disease-free periods ^8,9^. We believe that the limited availability of preventative or therapeutic options for recurrent aphthous stomatitis results from poor understanding of its etiopathogenic process ^10^, and the stagnation in our understanding of the condition over the last few decades. One of the biggest problems is that the scientific focus has been on studying individual molecules, such as TNF-α ^11^, IL-2 ^12^, and cortisol ^13^. In addition, much of the published research has not evaluated sample sets representing the clinical course of the disease. The sequence covers the stages of premonition (with symptoms but no visible signs of disease), pre-ulcerative (usually erythema and mild edema), ulcerative (acute or active phase, formation of the ulcer), healing (decrease in symptoms and progressive healing), and remission (no evidence of ulcers) ^14^. The ulcerative and remission phases are the stages able to be evaluated with greater objectivity in dental examinations ^15^.

Our research team wanted to try a different approach by applying state-of-the-art technology for the study of molecular processes: mass spectrometry (MS)-based proteomics. This technique analyzes a mixture of proteins, both qualitatively and quantitatively, using mass spectrometry. Additionally, the high feasibility and general sensitivity of this approach is particularly desirable for the identification and characterization of proteins ^16^. Proteomic technologies have a great capacity to generate a holistic view of complex processes that control health and disease states. To our knowledge, there are no studies applying global characterization of the proteome in patients with recurrent aphthous stomatitis.

Unfortunately, for research purposes, incisional or excisional biopsy of recurrent aphthous stomatitis ulcers is recommended only in cases of uncertainty ^17^. Saliva is an ideal source of biological information because it contains a set of biomolecules that cover oral structures, such as mucosa and teeth ^18^, and is therefore in close contact with ulcers. MS-based proteomics has been used to discover biomarkers in various oral conditions using saliva as a liquid biopsy, such as caries and periodontal disease ^19^, cleft palate ^20^, Sjögren’s syndrome ^21^, burning mouth syndrome ^22^ and oral cancer ^23^, but not in recurrent aphthous stomatitis. MS allows us to examine the salivary proteome in detail ^24^. Since saliva collection is less invasive and has no clinical complications associated with blood collection, substituting saliva samples for blood in biomarker analysis is of great interest ^25^.

Considering these antecedents, the objective of this study was to characterize the salivary proteome of patients with recurrent aphthous stomatitis using MS-based proteomics and assess its clinical usefulness in identifying the most representative biological and molecular processes during the course of lesions. To do this, we conducted a case-control study, analyzing the saliva of healthy controls and patients with recurrent aphthous stomatitis during a complete ulcerative cycle (active ulcers and absence of lesions). Using bottom-up proteomics, salivary proteins were sequenced with a timsTOF Pro mass spectrometer connected to nanoflow liquid chromatography (nLC-MS/MS). From these data, we were able to recognize biological processes related to vitamins, nutrients, interference of tissue destruction, and immune response against bacteria. We then identified bacterial proteins and the genera, species, and strains to which they belong. Finally, considering that recurrent aphthous stomatitis is tissue-specific, we evaluated the proteomic profiles of surfaces with and without ulcers. Applying a top-down proteomics approach, an Autoflex Speed MALDI-TOF mass spectrometer combined with machine learning analysis (MALDI-MS/ML) were used to obtain a specific and sensitive pattern of the salivary pellicle covering recurrent aphthous stomatitis ulcers and ulcer-free regions.

## METHODS

### General design

Using MS-based proteomics, we evaluated the salivary proteome of subjects with recurrent aphthous stomatitis in the presence and absence of ulcers. We also compared these profiles with healthy subjects without a history of aphthous ulceration. All procedures were performed in accordance with the Declaration of Helsinki. The Ethics Committee of the University of Antofagasta (protocol #156/2018) approved this research.

### Participants

We prospectively collected 118 salivary samples from 68 volunteers after obtaining informed consent. The subjects were divided into healthy controls (n=31; people with no history of oral ulcers) and recurrent aphthous stomatitis (n=36). The latter group was evaluated at study entry, during the ulcerative stage, and when lesions disappeared (remission stage, n=36). Additionally, we collected samples from recurrences that occurred after the remission phase (*n*=15). Subject demographics are provided in Supplementary Table 1. At the time of the examination, patients should have presented ulcers for no longer than 3 days. The exclusion criteria were the use of medications to treat ulcers during the previous 2 days and use of topical or systemic corticosteroids during the month before entering the study. Other exclusion criteria were the presence of other types of lesions of the oral mucosa, diseases with acute or chronic pain, smoking, excessive consumption of alcohol (more than three times weekly), and hematologic deficiency-related diseases. Patients with autoinflammatory syndromes, immunodeficiency states, gastrointestinal disorders, and hematinics deficits were also excluded (the complete list of diseases can be found in Supplementary Table S2). For their inclusion as controls, volunteers should never have presented oral ulcers, and except for the absence of lesions, this group shared the same inclusion and exclusion criteria.

### Salivary liquid biopsy

Subjects were requested not to eat or drink for at least 30 min before collection. All patients rinsed their mouths with 10 mL of water for 30 s. After 10 min, whole unstimulated salivary saliva collection began. Between 8:00 AM and 11:00 AM, each participant expectorated continuously in a 50 mL sterile propylene tube for a period of 5 min. Additionally, we collected the salivary pellicle deposited on lesions and the contralateral anatomical equivalent of the unaffected oral mucosa using cotton swabs. Immediately after collection, samples were deposited on ice and a protease inhibitor cocktail (cOmplete Tablets EASYpack, Roche) ^26^. We then centrifuged the samples at 14,000 × g at 4°C for 20 min to eliminate undissolved components and cellular detritus ^27^. The total protein concentration in the supernatants was determined using the BCA Protein Assay (Thermo Scientific). The samples were stored at −80°C for further processing.

### Protein identification and prioritization

When multiple subjects are being studied, pooling samples in proteomics experiments can help address resource constraints ^28^. Saliva samples were randomized and pooled into 9 sets: healthy controls (*n*=3, 11 samples per set), ulcerative (*n*=3, 12 samples per set), and remission phase (*n*=3, 12 samples per set). Total protein concentration for all samples was determined using the Pierce BCA Protein Assay kit (Thermo Scientific), following the manufacturer’s instructions for microplate procedures. Each sample contained 70 µg of protein. MS data were acquired by analyzing 500 ng of peptides after tryptic digestion and peptide fractionation using a nanoElute LC system coupled to a timsTOF Pro mass spectrometer (Bruker Daltonics). A detailed description of the instrument is provided in Refs. ^29^. MS raw files were processed using MsFragger through Fragpipe ^30^. For all searches, a protein sequence database of reviewed human proteins (accessed 06/15/2020 from UniProt; 74,823 entries) and bacterial proteins of species assigned to the oral cavity (accessed 12/11/2020 from The Human Microbiome Database; 728 files of annotated sequences from genomes) were used. Label-free protein quantification was performed using the IonQuant algorithm, as described previously ^30^. We used MSstats ^31^ to calculate protein abundance based on the ion abundances reported by MSFragger. We followed general workflow analysis for data-dependent acquisition (DDA or shotgun) experiments ^32^. The dataProcess function with log2 intensity transformation was used to calculate the protein abundance. We considered clinically significant salivary proteins with a log2 fold change value less than -0.5, or greater than 0.5, with a p-value ≤0.05, when comparing between groups (Student’s t-test). See Supplementary file (Methods section) for further details and explanation.

### Biological and molecular processes

To understand the set of clinically significant proteins, we performed an enrichment analysis. Pathway enrichment analysis helps to obtain a mechanistic view, identifying biological pathways that are enriched in a list of proteins more than would be expected by chance ^33^. To that end, we imported the proteins into the g:GOSt profiling online tool of G:Profiler ^34^ and queried for functional enrichment of terms derived from Kyoto Encyclopedia and WikiPathways. We used the g:SCS algorithm to address multiple testing, and a p-value ≤ 0.05 as a user-defined threshold for statistical significance. We also consulted the REACTOME database using the WebGestalt online tool ^35^. We considered those pathways with an FDR of ≤ 0.05. To understand the individual functions of each protein, we consulted QuickGO. With QuickGO, it is possible to determine a biological process or molecular event, with a known beginning and end, in which a protein participates ^36^.

### Western blotting, ELISA, and vitamin determination

We selected several proteins for the verification stage using western blotting and ELISA. Briefly, 8□μg of total salivary protein was subjected to SDS-PAGE on a 4%–20% precast gel (Bio-Rad #456-1093). Western blotting was subsequently performed using anti-CA1 (1:450, Abcam #ab182609) and anti-SLURP1 (1 µg/mL, Abcam #ab3840) as primary antibodies. After incubation with a secondary anti-rabbit antibody (1:5,000, Abcam #ab97048), proteins were visualized using alkaline phosphatase chromogen (NBT/BCIP, Abcam #7468). Ponceau red was used as the loading control. The uncropped scans of the gels and blots are shown in Supplementary Figure 1. ELISAs were performed using 13□μg of total salivary protein as the sample with HBB (Abcam #ab235654), CST1 (Sigma–Aldrich #RAB1036), and A2M (Sigma–Aldrich #RAB0600) kits, according to the manufacturer’s instructions. Folate (range: 0.35–24 ng/mL) and vitamin B12 (range: 45–2,000 pg/mL) concentrations were determined on an ADVIA Centaur XPT immunoassay system (Siemens) according to the manufacturer’s instructions.

### Proteomic profiles comparison

We analyzed the affected and unaffected oral mucosa using cotton swab samples. Given its speed, simplicity, and low cost, as well as the availability of equipment and expertise in many hospital laboratories in developing countries, MALDI-MS/ML is a promising alternative ^37^. In this experiment, we did not pool samples (each was analyzed individually) and we added new samples from ulcer recurrences (new lesions after the remission stage) experienced by some of the patients (n=15). Samples were analyzed on an Autoflex Speed MALDI-TOF mass spectrometer (Bruker Daltonics) following the protocol recently published by our group ^37^ and detailed in Supplementary File (methods section). Proteomic profiles were obtained and then preprocessed to generate a matrix of peak intensities. The most relevant peaks were selected using the correlation-based feature subset selection method (CFS) ^38^ implemented in the Weka software ^39^. To explore and compare spectra in multidimensional space, principal component analysis (PCA) was performed using the R packages FactoMineR ^40^ and factoextra ^41^ (data were scaled to unit variance).

### Sample Size Calculation

The number of patients was estimated following guidelines for the use of MS-based proteomics in clinical studies ^42,43^. We consider a two-sided hypothesis test with a confidence level of 99.9% (α=0.001) and a power of 95% (1-β = 0.95). Changes in protein abundance or effect size (relative intensity data) were considered as differences based on log2 (0.5). Biological variation (containing technical variation) was established with a magnitude of 0.5. We performed each analysis once. The number of participants was adjusted to a probable loss of 20%, establishing a minimum of 31 volunteers per group.

## RESULTS

### Clinicopathological characteristics

The subjects were from the same population base. The mean age of the participants was 26 years for both groups, with the majority being women. The profile of our population agrees with that reported in the international literature, both in age and in the predisposition for the appearance of these lesions in women ^44,45^. Figure 1 shows representative ulcers observed in this research. Most lesions occurred in the non-keratinized oral mucosa, where the lips were the most affected territory (Supplementary Table 1). Ulcers mainly corresponded to single lesions classified as minor, with moderate pain according to the visual analog scale. The participants reported presenting more than seven episodes per year. Considering that recurrent aphthous stomatitis lasts for approximately two weeks, the large number of ulcer events highlights the clinical relevance of the condition.

**Figure 1.**
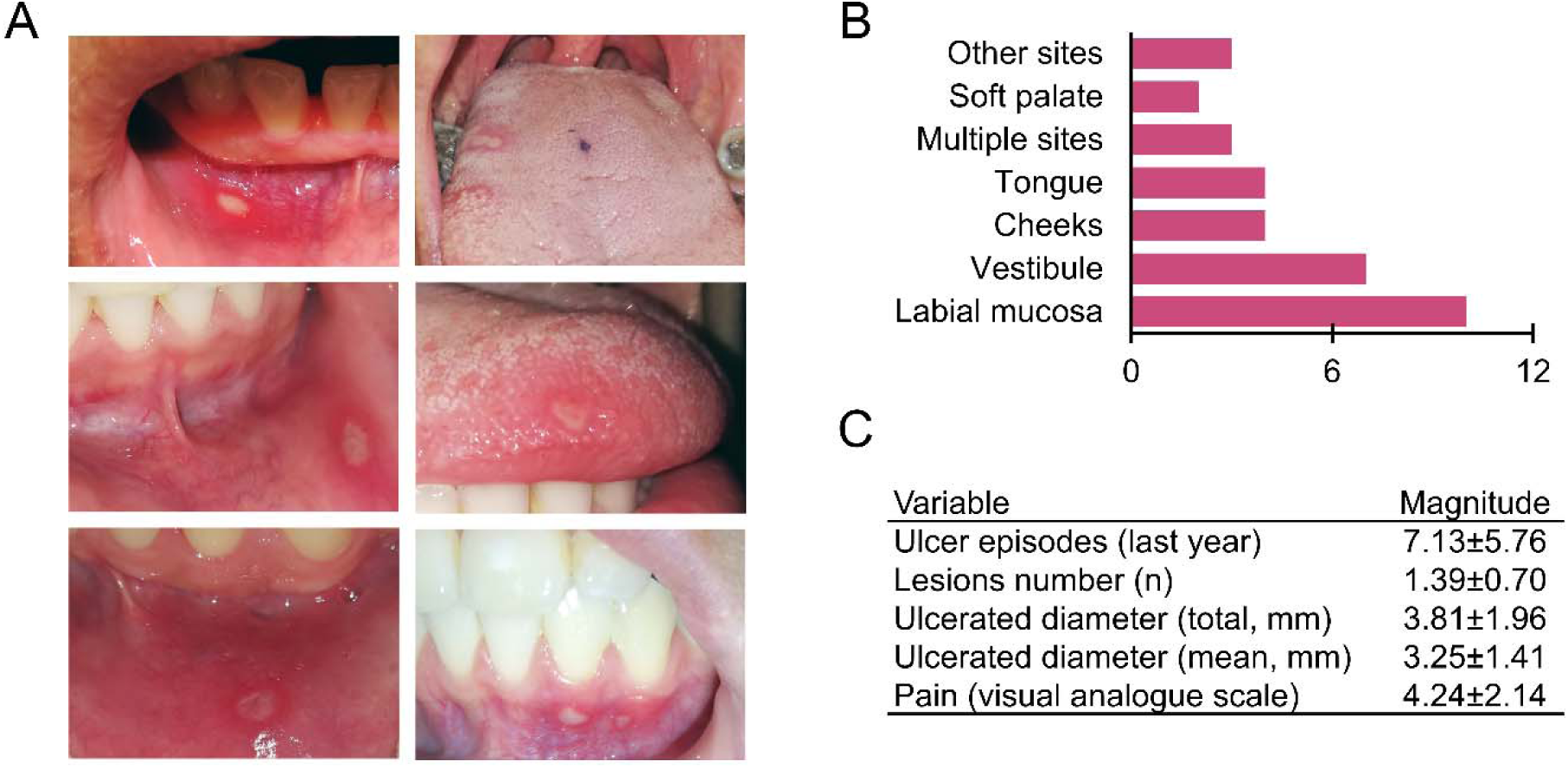
Clinical manifestations of recurrent aphthous stomatitis. (**A**) Representative images of ulcers. Lesions were ovoid with an erythematous halo. (**B**) Anatomical sites where lesions occurred. The most affected regions were labial mucosa, vestibule, and cheeks. **(C**) Clinical history associated with ulcers. Lesions were smaller than 5 mm accompanied by moderate pain.

### Proteomic analysis revealed the differentially expressed proteins

To identify salivary proteins in the health, ulcerative, and remission stages, we prepared three sets of pooled samples (pools), each containing one-third of the subjects. With a flow that included nLC-MS/MS associated with the Fragpipe tool, we were able to identify more than 1,200 proteins in each set (Figure 2B). In healthy controls, we found 10 unique proteins (HABP2, HEBP1, HTRA1, ILF2, LDLR, MMP7, PADI3, PTBP1, PTN6, and RALB), 5 in the ulcerative stage (APOL1, C1QBP, FHR2, HV124, and NQO2), and 8 in the remission stage (BAX, BIN2, CTNA1, DP13A, FA20C, PP14B, RS20, and TBB4A). Pooling may explain the low number of unique proteins. Therefore, although there were exclusive proteins in each group, we were interested in those that were in common. We assume that comparative analysis allows for a better understanding of the biological context. For this, we concentrated on more than a thousand proteins shared by all the groups and that, according to the MSStats flow, presented a relevant fold change (blue and red points in volcano graphs, Figure 2C).

**Figure 2.**
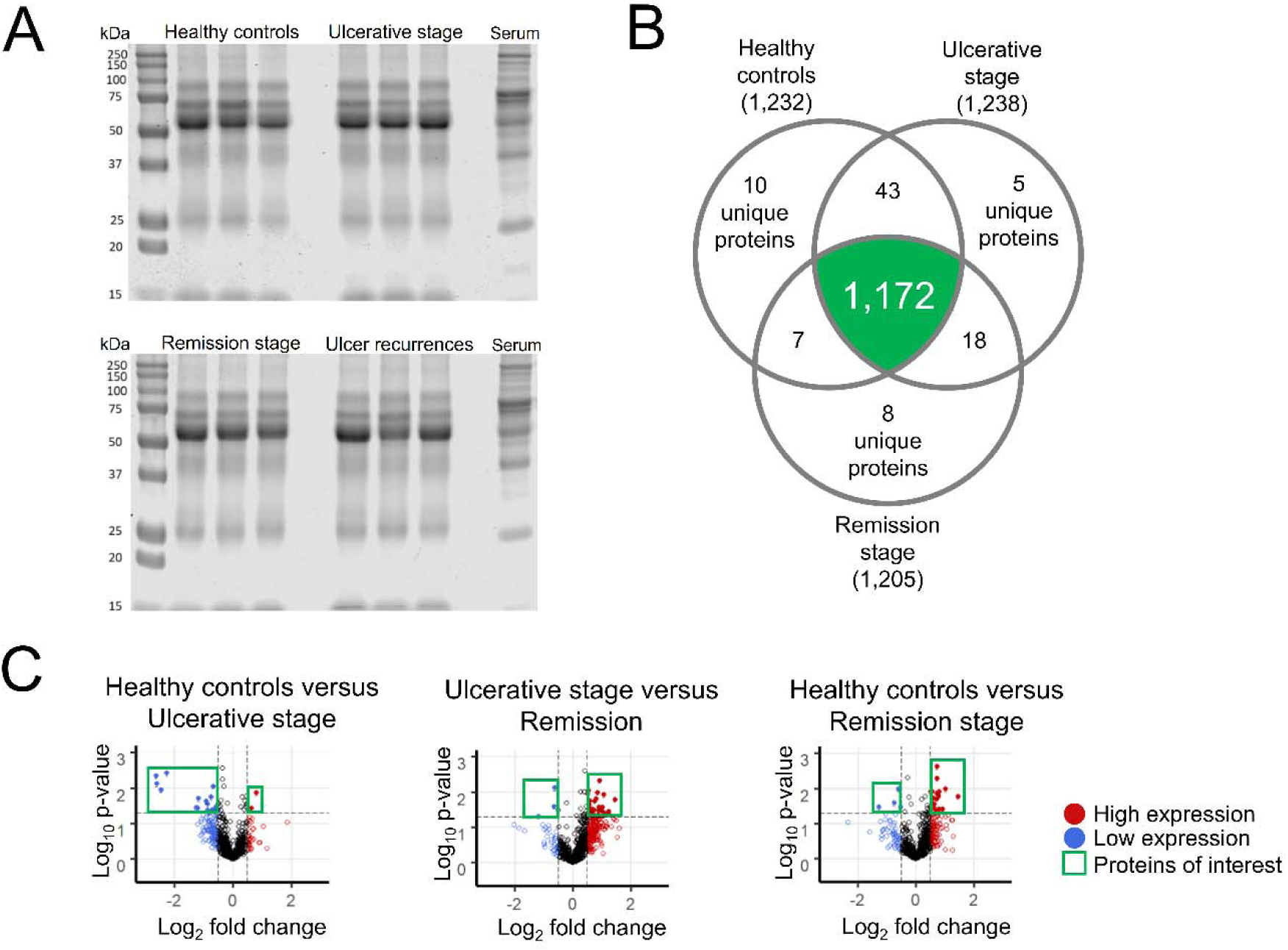
Salivary proteome. (**A**) We assessed samples integrity by SDS-PAGE gel electrophoresis (20 μg/lane protein concentration). (**B**) Venn diagram shows that most of the salivary proteins identified were proteins in common. (**C**) Volcano graphs show the windows considered in the analysis, constructed considering statistical significance and fold change. Proteins up- and down- regulated are enclosed by a green square. The fold change limits were -0.5 and 0.5. The established p-value was 0.05.

### The presence and absence of lesions are associated with pathways related to vitamins and nutrients

To understand the context in which the protein collective might participate, we performed an enrichment analysis using g-Profiler. For this, we included all differentially expressed proteins. Figure 3 shows the comparison between the healthy control group and the ulcerative stage patients (Figure 3A) and the ulcerative stage versus the remission-stage patients (Figure 3B). These comparisons revealed the metabolic pathways of macronutrients and micronutrients (nitrogen and selenium) along with the digestion and absorption of vitamins (B9 and B12). Notably, there was no enrichment when analyzing the list of differentially expressed proteins between healthy control samples and the remission stage. In other words, in terms of signaling, there were no differences between the groups. This result indicates that alterations in signaling pathways are only present during the clinical manifestation of the ulcer. It is necessary to note that the enrichment analysis placed a part of the identified proteins into context; therefore, to complete the picture, it is also necessary to describe the individual biological roles of each of them.

**Figure 3.**
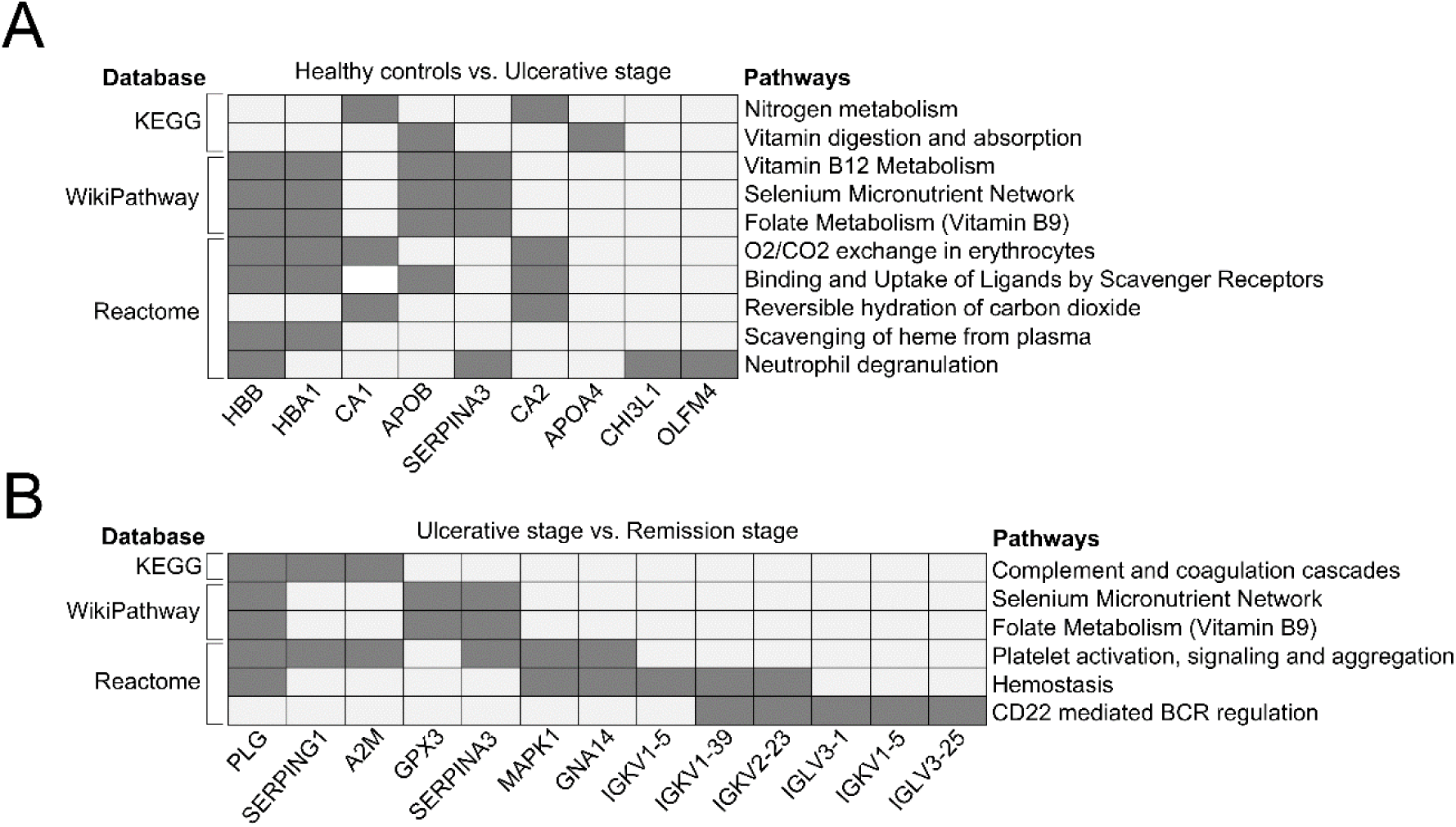
The presence or absence of recurrent aphthous ulcers reveals metabolic pathways for vitamins and nutrients. Each pathway on the right corresponds to a database on the left. The existence of proteins in these pathways is indicated by dark gray squares. APOB and SERPINA3, for example, are proteins involved in the vitamin B9 and B12 metabolism pathways. (A) Pathways enriched from the list of differentially expressed proteins between healthy controls and patients at the ulcerative stage. (B) Enrichment analysis corresponding to the contrast between the ulcerative and remission-stage patients. The absence of oral ulcers, i.e., the contrast between healthy controls and remission-stage patients, did not show enriched pathways. For enrichment analysis, g:Profiler (https://biit.cs.ut.ee/gprofiler/gost) and WebGestalt (http://www.webgestalt.org/) were used.

### Proteins involved in the inhibition of proteolysis and responses against bacteria stand out in the presence of lesions

Because the enrichment analysis did not provide information on all proteins, we list each differentially expressed protein in Table 1. To understand their individual roles, we examined the functional annotations in QuickGO. In contrast, information from patients with recurrent aphthous stomatitis during the ulcerative stage and remission stands out. Several upregulated proteins participate in the negative regulation of peptidase activity (SERPINA6, SERPING1, SERPINA3, and A2M) and the immune response against bacteria (IGHV1-8, IGHV1-69D, IGHV3-74, IGHV4-30-2, IGHV3 -43, IGHV3-66, IGHV3-16). This result suggests that the ulcerative activity may be a response to the presence of a microorganism that is harmful to the oral mucosa or that the presence of lesions alters the microbiota and that a decrease in proteolysis is necessary for the regeneration of lesions.

**Table 1.**
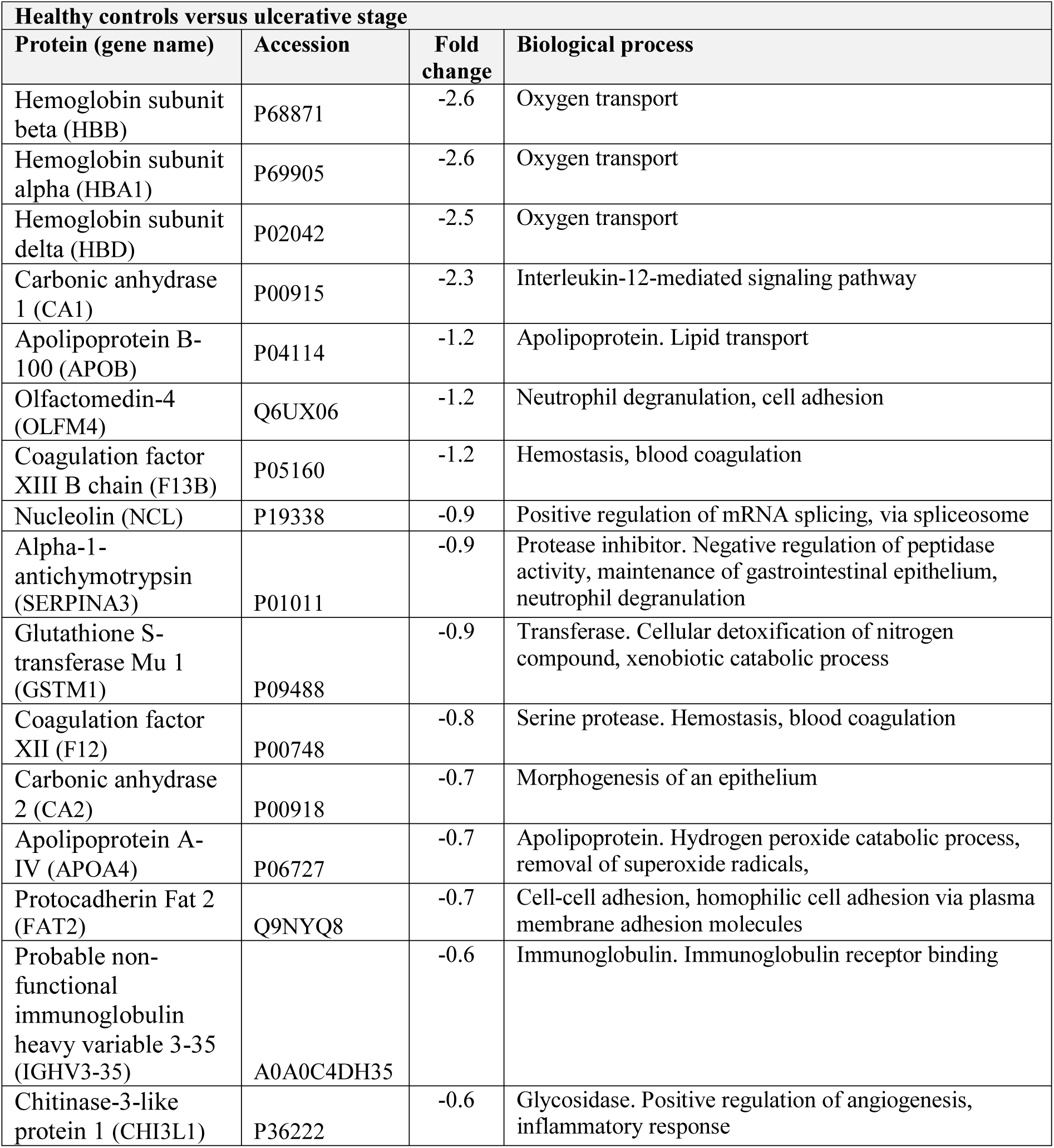
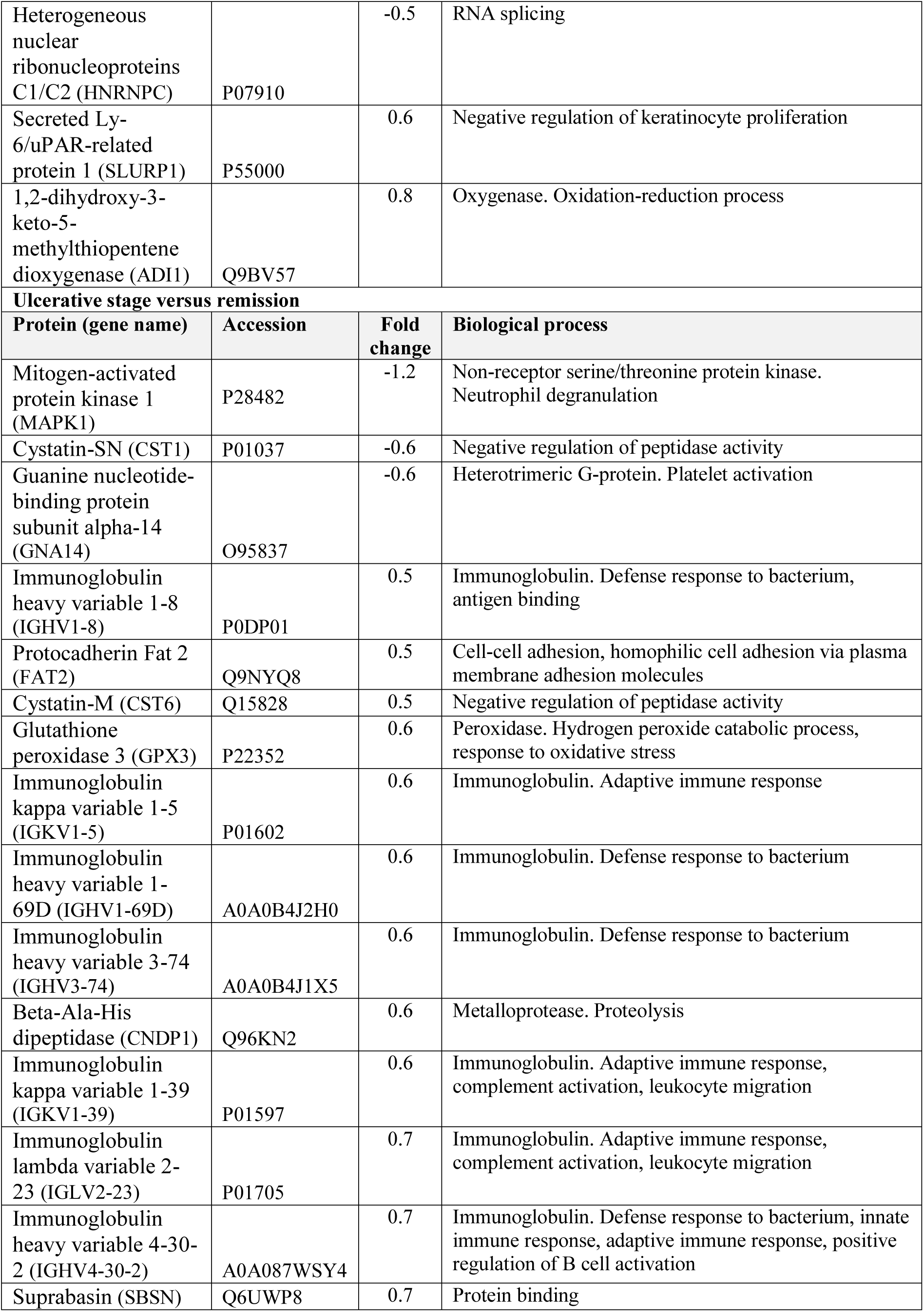
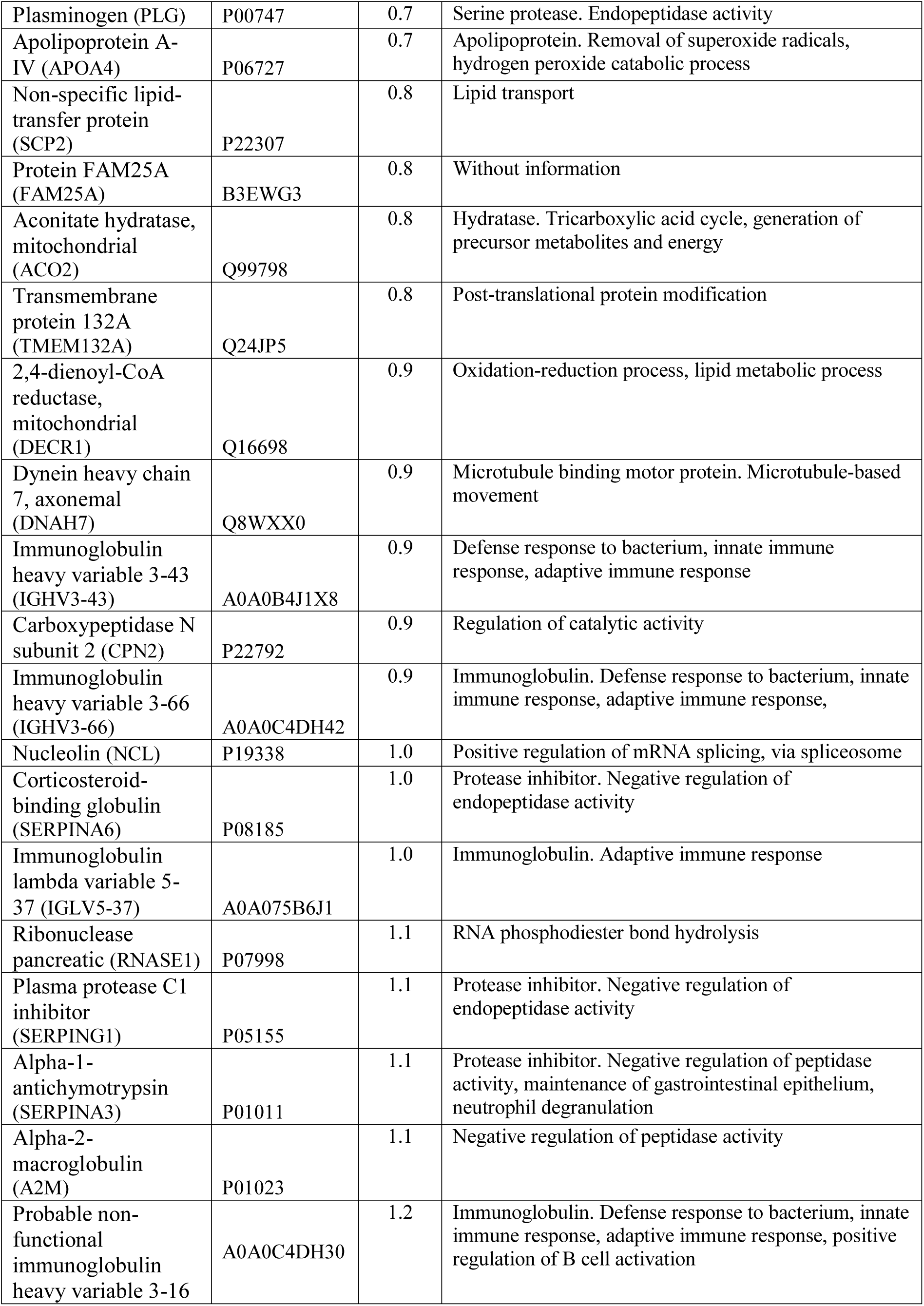
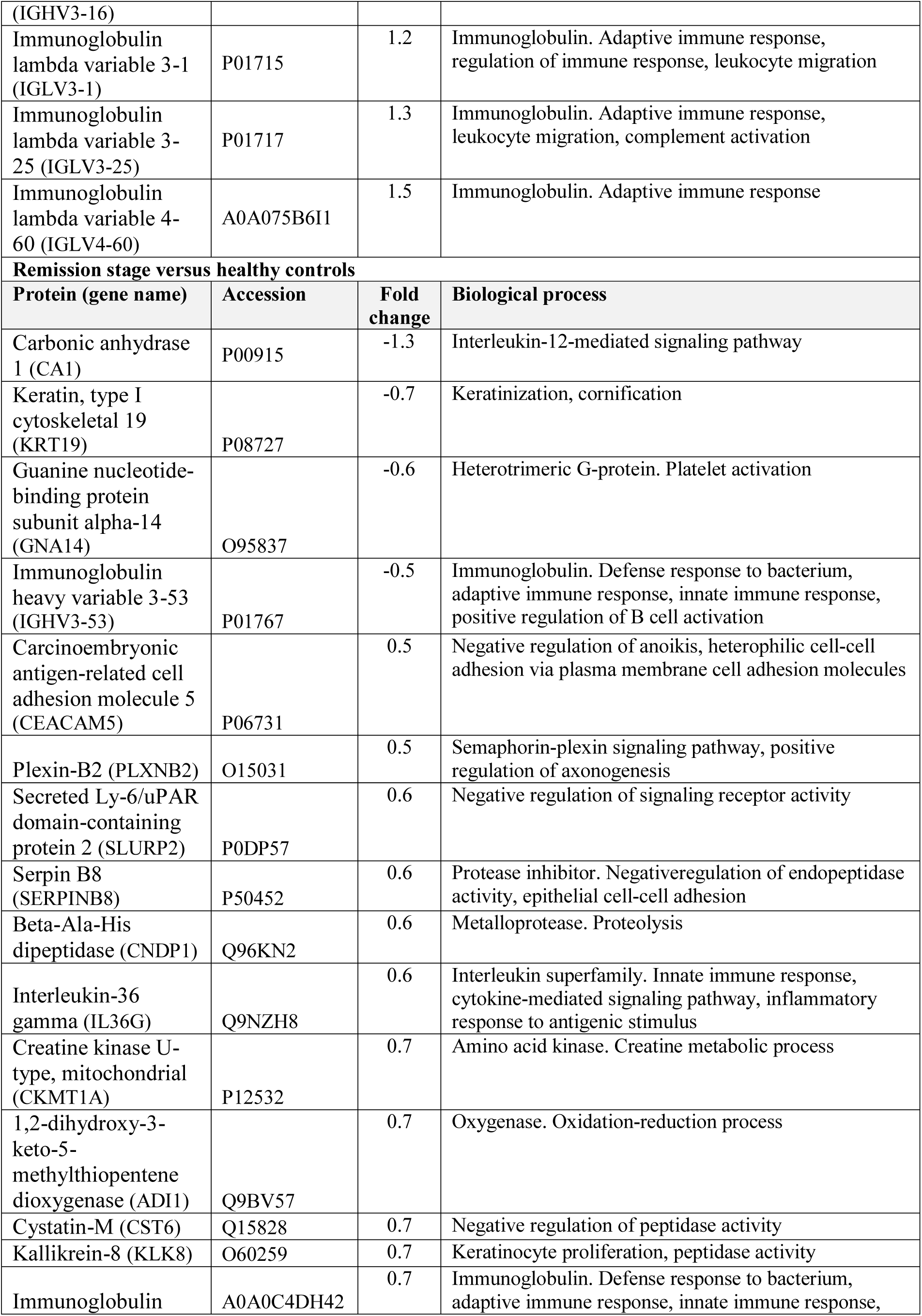
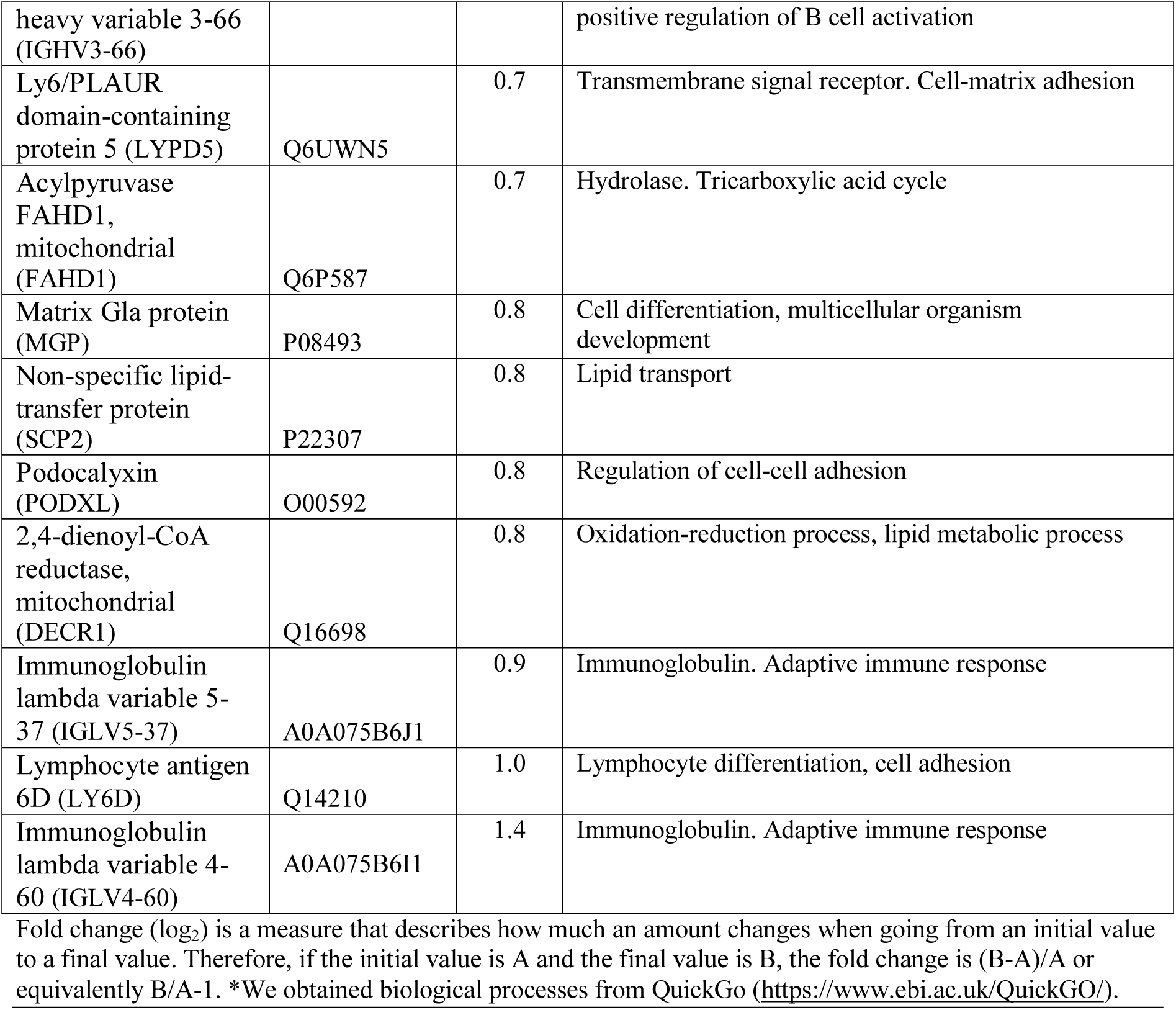
Biological processes of differentially expressed proteins in recurrent aphthous stomatitis.

### Proteomic data verification

To validate the data points, we used some of the most extreme fold-change values. Results of western blotting and ELISA confirmed the presence of CA1, SLURP1, HBB, CST1, and A2M in the analyzed saliva pools (Figure 4). In this orthogonal validation, CA1 and HBB levels were elevated in recurrent aphthous stomatitis (ulcerative and remission stages, Figure 4A and B upper panels, respectively). Additionally, we checked for the presence of vitamins B9 and B12 in the saliva. In general, patients with recurrent aphthous stomatitis had lower vitamin levels compared to controls but without significant differences (Figure 4C).

**Figure 4.**
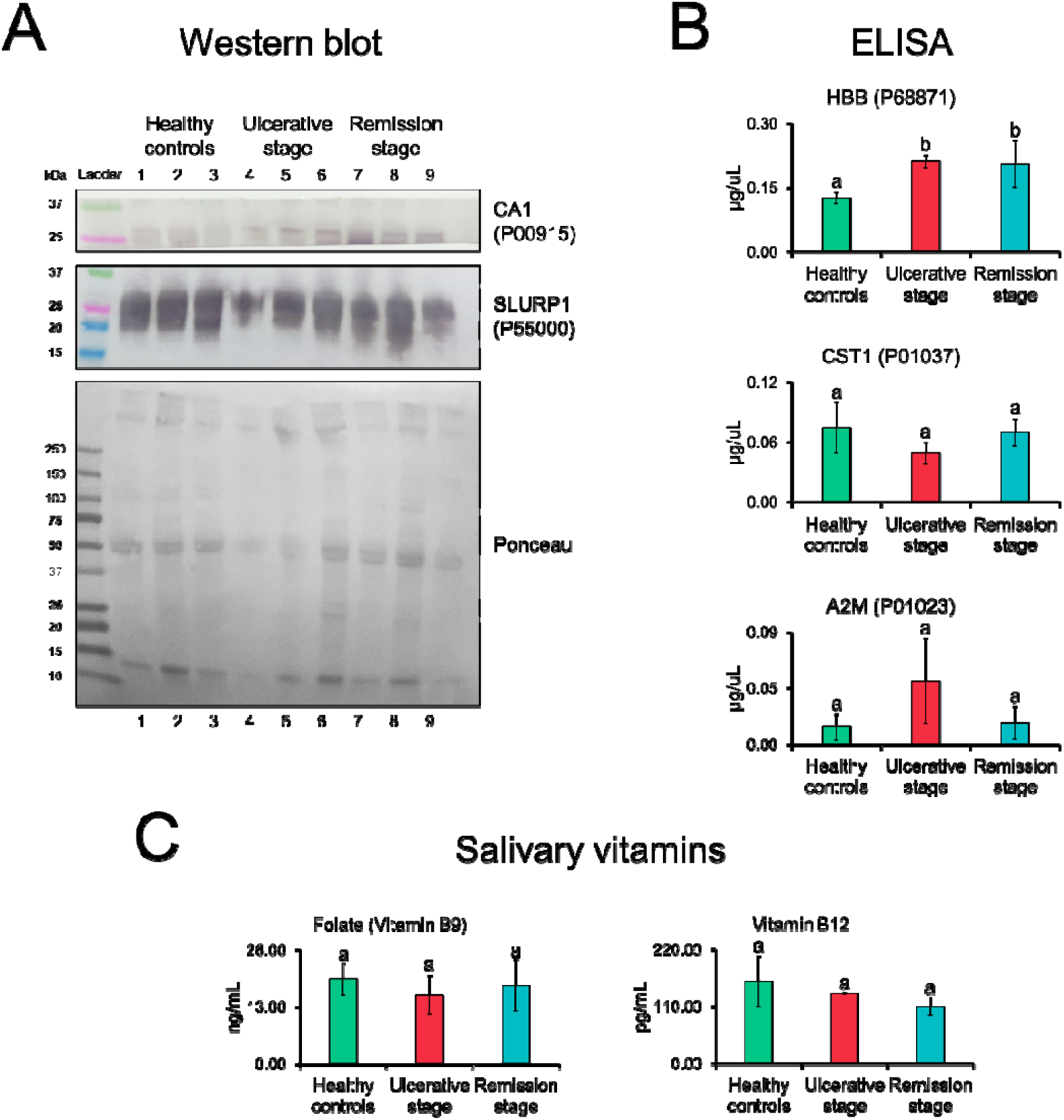
Salivary proteins and vitamins during the clinical course of recurrent aphthous stomatitis. Verification of proteomic data points. (**A**) Western blotting showed higher CA1 protein levels during the ulcerative and remission stages. Ponceau red were used as loading control. (**B**) ELISA showed a higher amount of HBB protein in ulcerative and remission stages. (**C**) Samples from patients with recurrent aphthous stomatitis show lower levels of folate (vitamin B9) and vitamin B12, but without statistically significant differences. Data are represented by the mean□±□SD (Student’s t-test, different letters indicate statistically difference at p-value□≤□0.05). CA1, carbonic anhydrase 1; SLURP1, secreted Ly-6/uPAR-related protein 1; HBB, hemoglobin subunit beta; CST1, cystatin-SN; A2M, alpha-2-macroglobulin.

### *Neisseria meningitidis* proteins were present at the ulcerative stage

Because defense against bacteria was one of the prominent pathways identified, we investigated whether we could detect bacterial proteins and species in salivary proteomes. For this purpose, we downloaded all the protein sequences of the bacterial species assigned to the oral cavity in the Human Oral Microbiome Database ^46,47^. The resulting database was approximately 46 times larger than that of humans (1,174,712 KB vs. 26,318 KB). We identified 134 bacterial proteins. Figure 5 shows that two proteins, isocitrate dehydrogenase [NADP] (icd, UniProtKB Q9JZS1) and elongation factor Ts (tsf, UniProtKB P64051), belonging to *Neisseria meningitidis* strains were elevated only in the presence of lesions. The response indicated by the salivary proteome against bacteria could result from the activity of *Neisseria meningitidis*.

**Figure 5.**
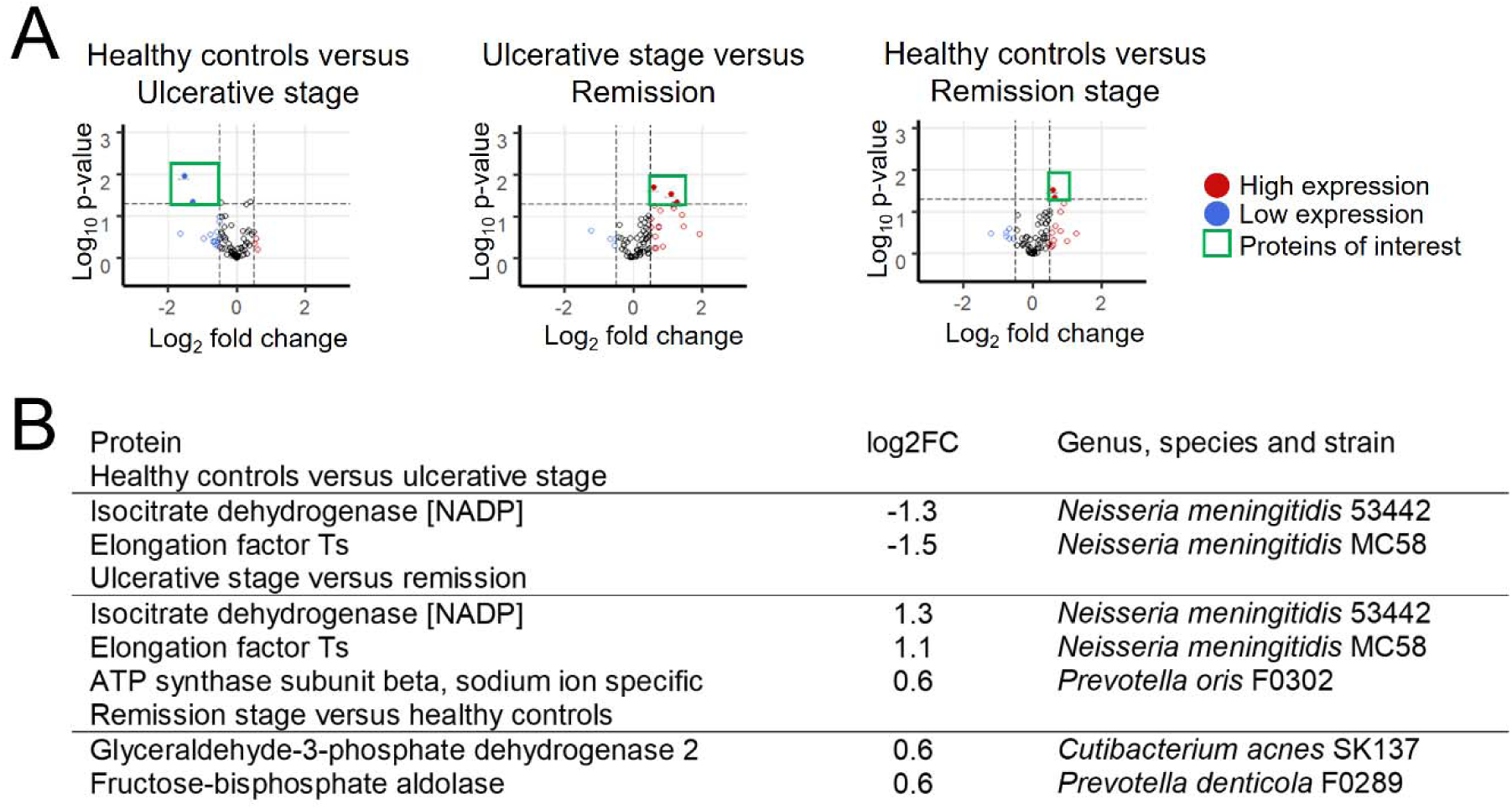
Presence of recurrent aphthous ulcers reveals *Neisseria meningitidis* proteins. (**a**) Volcano graphs show the windows considered in the analysis, constructed considering statistical significance and fold change. Proteins up- and down-regulated are enclosed by a green square. Fold change limits were -0.5 and 0.5. The established p-value was 0.05. (**b**) The table details the proteins expressed differentially in the comparison between the groups. The presence of proteins and strains of *Neisseria meningitidis* (serogroup B, strain MC58 and serogroup C, strain 053442) during ulcerative activity stands out. Information of oral bacteria was obtained from The Human Oral Microbiome Database (http://www.homd.org/).

### The oral surface epithelium without lesions did not show any differences between the groups

As recurrent aphthous stomatitis is a tissue-specific condition, we analyzed the salivary pellicle that covered the area affected by recurrent aphthous ulcers and the healthy contralateral anatomical region, to provide a local picture. In this analysis, we also included recurrent lesions (new ulcerative stages after remission). MALDI-MS/ML analysis showed that the proteomic profiles (mass spectra) were different when presence and absence of lesions were compared in a multidimensional space (PCA graphs, Figure 6). However, the data from the healthy tissue regions could not be separated completely, both at the remission stage (Figure 6b) and at all unaffected oral mucosa sites (Figure 5d). This evidence shows that the salivary pellicle that covers the epithelium differs only during active lesions, but not during epithelial integrity.

**Figure 6.**
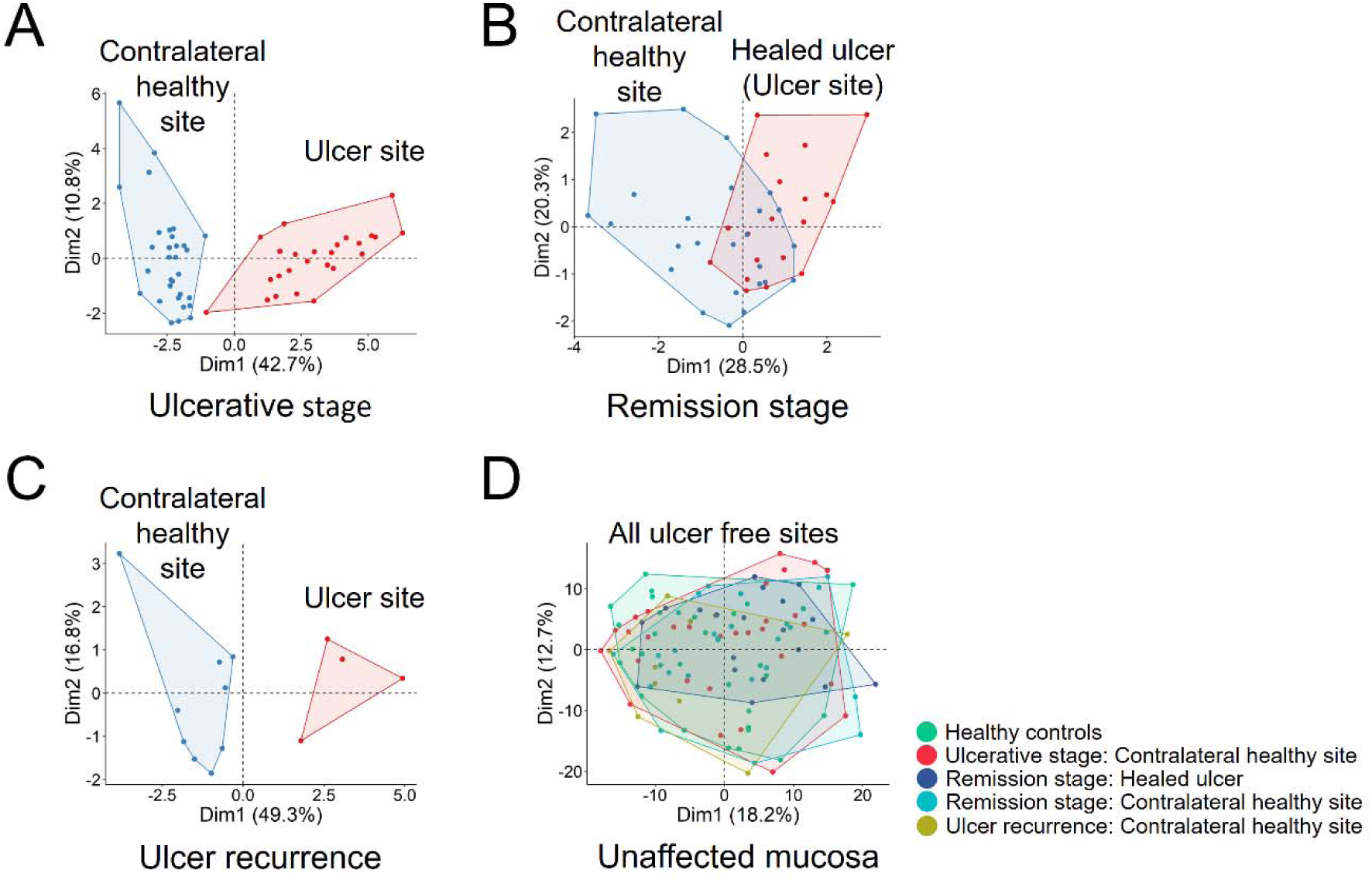
Recurrent aphthous stomatitis did not leave a mark on affected sites. Selected peaks with CFS (see table S4 in supplementary material) from proteomic profiles were used to perform PCA graphics. (**A**) During the ulcerative stage, healthy and affected tissues are completely separated. (**B**) As the oral mucosa regenerates, healthy and recovered tissues fail to separate. (**C**) When an injury occurs again, the sites separate again. (**D**) All intact oral mucosa tissues fail to separate, regardless of health and disease status.

## DISCUSSION

In this investigation, we determined the salivary proteome of patients with recurrent aphthous stomatitis during the presence and absence of lesions using proteomics based on mass spectrometry. Our analyses revealed that the ulcerative stage is related to several biological pathways, including vitamin and nutrient metabolism, inhibition of tissue destruction, and defense against bacteria, probably *Neisseria meningitidis*. Additionally, we established that the proteomic profiles of ulcer-free states do not differ from those of healthy individuals. Our evidence suggests that recurrent aphthous stomatitis corresponds to an oral manifestation of a temporary local or systemic condition.

Because tissue biopsy is not indicated in recurrent aphthous stomatitis ^15^, we used saliva as a “liquid biopsy” to obtain information on the health status of the participants. The lesions corresponded to small, solitary or multiple ulcers, with moderate pain, present almost exclusively in the non-keratinized oral mucosa. These characteristics are consistent with the classic profile of recurrent aphthous stomatitis ^48,49^. In the analysis flow with nLC-MS/MS and bioinformatics, we used the fold change as a measure of contrast between the groups. From these values, we concentrated on the salivary proteins that presented a differential expression on which we explored the biological information. Fold change is a value that reports the extent to which a quantity changes from one state to another. It is used in proteomics to evaluate the differential expression of proteins ^50^ and to identify disease biomarkers ^51^.

Through enrichment analysis with g:Profiler and WebGestalt, we found that ulcerative activity is related to the metabolism of vitamins (B9 and B12) and nutrients (selenium and nitrogen). Our evidence complements one of the main lines of inquiry for explaining the etiopathogenesis and severity of recurrent aphthous stomatitis with respect to nutritional deficiencies; low blood levels of iron ^52^, vitamin D ^53-55^, vitamin B9 ^56^ and B12 ^57,58^ have previously been reported to be associated with the presence of lesions. In our study, the levels of salivary vitamins B9 and B12 did not show significant differences. This may be due to the fact that there is a low correlation between serum and salivary levels, as has already been demonstrated in the geographic tongue ^59^. Some of hematinic deficiencies already named can lead to anemia in patients with recurrent aphthous stomatitis ^60^. Previous studies have shown that anemia causes atrophy of the oral epithelium, leading to a decrease in the mucosal barrier ^61^. Selenium deficiency can also make the host susceptible to infection ^62^. The presence of these molecules was not examined in our patients, as it was outside the objectives of our research; this may constitute a limitation in our analyses, and it would be advisable to measure these molecules in the future. However, it is interesting that saliva represents, at least indirectly, the nutritional states previously described in the literature in these patients. The high expression of salivary proteins associated with metabolic pathways may be attributed to the lack of vitamins and nutrients and suggests that these proteins may not be involved in these pathways (i.e., they are freely available). In this sense, it has been shown that supplementation with vitamin B12 decreases the duration of ulcer episodes, number of injuries, and pain ^63^. The finding of no differences between healthy subjects and those with recurrent aphthous stomatitis in the remission stage was particularly notable, as it indicates that these two clinical states are not significantly different; alterations that determine an ulcerative state are silenced in the absence of lesions. This evidence indicates that diet plays an important role in triggering lesions and that the aforementioned alterations may be temporary and susceptible to intervention.

The enrichment analysis showed a high level of various proteins during the ulcerative stage. Interestingly, HBB and SERPINA3 have anti-inflammatory effects. Using ELISA, we verified that patients with recurrent aphthous stomatitis present a high expression of HBB. HBB can be cleaved into two chains, one of which is called spinorphin. Spinorphin inhibits various neutrophil functions ^64^, and additionally promotes pain reduction by inhibiting the degradation of endogenous opioids ^65^. SERPINA3 inhibits neutrophil cathepsin G protease, a pro-inflammatory enzyme released at sites of inflammation that contributes to the activation of inflammatory cytokines, pathogen degradation, and tissue remodeling ^66^. Inhibition of cathepsin G likely contributes to reduction of tissue damage from proteases that are released at the site of epithelial injury ^67^. Repeated topical application of SERPINA3 (2 mg/mL) on induced wounds in mice has been found to significantly accelerated their closure ^67^. Considering that the enrichment analysis did not provide information on all proteins, we consulted each of them individually in QuickGo. In this analysis, a protective response stood out again, and peptidase activity was negatively regulated (SERPINA6, SERPING1, SERPINA3, and A2M). In a recent study, Bao et al. used LC-MS/MS and miRNA microarrays to evaluate the serum from patients suffering from recurrent aphthous ulcers ^68^. The results showed SERPINA1 upregulation in patients with lesions compared to that in healthy controls. Serpins are protease inhibitor proteins that inhibit target enzymes by causing conformational changes. Many key proteolytic cascades, including mammalian coagulation processes, are regulated by these proteins. Together, these data indicate that after epithelial destruction, proteins are released to inhibit tissue destruction promoted by neutrophils. ^69^.

In our experiments we verified a high expression of CA1 during the ulcerative and remission stages. CA1 is a member of the CA family of carbonic anhydrases, which reversibly catalyzes the hydration of CO_2_ to generate HCO_3_−, which then rapidly binds to calcium ions to produce calcium carbonate ^70^. Carbonic anhydrases are known to modulate immune cell activation. Evidence from experiments in mice shows that inhibition of CA1 possesses therapeutic potential that can be used to treat mast cell-mediated inflammation ^71^. Pre-ulcerative lesion of recurrent aphthous stomatitis demonstrates subepithelial inflammatory mononuclear cells with abundant mast cells ^72^. In addition, it has been reported that the presence of mast cells and their degranulation activity at the connective tissue of recurrent aphthous stomatitis lesions does not necessarily mean that they play a destructive role; mast cells also have the ability to mediate a protective role and might be involved in the repair process ^73^. Whether in a destructive or protective sense, salivary CA1 could be related in some way to those activities. Subjects with ulcers therefore have active processes to destroy, or conversely, recover and preserve the integrity of the oral mucosa toward the remission stage. These processes have been induced by, or induce changes to, the salivary proteome.

Another relevant biological process identified was the immune response against bacteria, associated with several differentially expressed proteins (IGHV1-8, IGHV1-69D, IGHV3-74, IGHV4-30-2, IGHV3-43, IGHV3-66, IGHV3-16). To further investigate these findings, we analyzed over 700 species sequence files from the oral microbiome. Although the samples were centrifuged, we were able to identify that two strains of *Neisseria meningitidis* (53442 and MC58) were consistently present only in the ulcerative stage when compared to states without lesions. This is due to the identification of proteins belonging to this bacterium (isocitrate dehydrogenase [NADP] and elongation factor Ts). Isocitrate dehydrogenase [NADP] participates in the synthesis of NADPH, which provides reducing power that drives numerous anabolic reactions, including those responsible for the biosynthesis of all major cellular components ^74^. The elongation factor Ts participates in the elongation of the polypeptide chain during prokaryotic translation, allowing the synthesis of proteins ^75^. This leads us to infer that *Neisseria meningitidis* can be found in an active phase of growth and synthesis. This result should be interpreted with caution because bacteria in the oral cavity may exhibit considerable sequence similarity ^76^. *Neisseria meningitidis* is a transient commensal of the human oropharynx that causes life-threatening diseases, such as meningitis and bacteremia ^77^. The asymptomatic oropharyngeal carriage of *Neisseria meningitidis* reaches its peak in adolescents and young adults ^78^. This is consistent with the peak and frequency of recurrent aphthous stomatitis in the general population ^48,79,80^. A recent metagenomic study showed that *Neisseria meningitidis* is correlated with several oral bacterial species, including *Fusobacterium nucleatum, Aggregatibacter aphrophilus, Campylobacter rectus, Catonella morbi, Haemophilus haemolyticus*, and *Parvimonas micra* ^77^. The researchers suggest the ecological interactions of *Neisseria meningitidis* with the oral microbiota extend throughout the oral cavity. This result ties into another dominant line of inquiry seeking to explain the cause of recurrent aphthous stomatitis: changes in the oral microbiome. The alteration of the normal oral microbiota triggers the presence of lesions, and the presence of lesions alters the microbiota ^81^, even after the lesions have healed ^82^. There are several studies that employed pyrosequencing of 16S rRNA genes to demonstrate dysbiosis with different bacteria, including *Streptococcus salivarius* and *Acinetobacter johnsonii* ^83^, *Escherichia coli* ^84^ and bacteria of the phylum Bacteroidetes ^84,85^. The use of mass spectrometry-based proteomics to identify proteins in saliva is reliable. A previous study showed that the percentage of tryptic peptides that share the same sequence between humans and bacteria is only 0.04%^86^. Our findings are limited to salivary identification and should subsequently be subsequently verified in microbiological cultures from salivary samples of patients with recurrent aphthous stomatitis in different phases of the disease to elucidate potential causality. Furthermore, all the biological knowledge consolidating our arguments has been sourced from public databases, whose information evolves over time. Considering that the saliva was centrifuged, a good biological resource to exploit further is the pellet; from this, PCR or MALDI Biotyper could be used to explore the presence of *Neisseria meningitidis* and other bacteria for complete understanding of an eventual oral dysbiosis.

Our results show that, from a systemic point of view, the saliva of subjects who had never presented ulcers compared to subjects with recurrent aphthous stomatitis in the remission stage did not present differences in terms of biological processes. Considering that this condition is tissue specific, we examined the adhered salivary pellicle on the surfaces with and without lesions using MALDI-MS/ML. In this last experiment, we also added some samples of ulcer recurrence that patients made after the remission stage. MALDI-MS/ML analysis showed that the proteomic profiles of all lesion-free states did not differ, except when the lesion was present. This emphasizes that if recurrent aphthous stomatitis is considered a disease, it does not leave a mark on the epithelial salivary pellicle. Furthermore, when comparing the lesion-free states, no enriched biological processes are found, leading us to reflect on the true nature of recurrent aphthous stomatitis. The activity of recurrent aphthous stomatitis represents a pathological condition; according to the Medical Subject Headings, pathological conditions are abnormal anatomical or physiological conditions and objective or subjective manifestations of the disease, not classified as a disease or syndrome (MeSH Unique ID: D013568). There are various diagnoses and systemic states that occur with aphthous ulcerations, such as PFAPA syndrome, Behçet’s syndrome, Crohn’s disease, celiac disease, HIV infection, clinical neutropenia, among several others. There are many paths for the same lesion. Our data and results lead us to hypothesize that recurrent aphthous stomatitis is an oral manifestation on a local or systemic basis in susceptible subjects. This is likely due to nutritional deficiencies and bacteria-triggered lesions normally found in the oral cavity of adolescents and young adults.

In this investigation, data obtained by mass spectrometry-based proteomics cumulatively supported the role of vitamin metabolism, nutrients, and bacteria in recurrent aphthous stomatitis. We demonstrated the usefulness of salivary biopsy for obtaining systemic and local information. From a clinical perspective, we specifically suggest the participation of proteins involved in the metabolism of vitamins B9 and B12, the nutrients nitrogen and selenium, and the activity of *Neisseria meningitidis*. Our evidence deepens the existing knowledge on the etiopathogenesis of recurrent aphthous stomatitis, using an approach that has never been utilized in this context. The conceptual framework that we have established here should be confirmed with subsequent experiments that seek, first, to define a state of systemic susceptibility, which would allow the establishment of lesions in the presence of an etiological candidate agent. Subsequent research may enable the development of preventative approaches for this disease, which are currently unable to be achieved.

## Supporting information

Database

## Data Availability

All supplementary files are included in this article and are publicly available at: https://doi.org/10.5281/zenodo.4482898. The mass spectrometry proteomic data have been deposited in the ProteomeXchange Consortium via the PRIDE partner repository 87 with the dataset identifier PXD026401 (https://www.ebi.ac.uk/pride/archive/projects/PXD026401). All other data supporting the findings of this study are available from the corresponding author on reasonable request.

https://www.ebi.ac.uk/pride/archive/projects/PXD026401

## Acknowledgments

We want to thank all the volunteers who actively participated in this research, despite the national social context and the global pandemic, and the University Dental Clinic Center for hosting us. We also want to give special recognition to the TENS Yennifer Lemus. We would like to express our gratitude to Fengchao Yu (Department of Pathology, University of Michigan, Ann Arbor, Michigan, USA) for his assistance with the Fragpipe. We acknowledge Melisa Institute (San Pedro de la Paz, Chile), Laboratory of Asymmetric Synthesis (Talca, Chile), and Loncomilla Laboratory (Talca, Chile) for providing the infrastructure and human resources to perform crucial experiments. Funding was provided by the Chilean National Agency for Research and Development (ANID) PhD grant #2016-21161314 (to A.P.), ANID FONDECYT Regular num. #1180084 (to F.M.N. and L.S.S.), ANID FONDECYT Iniciación num. #11180170 (to C.R.) and Concurso de Proyectos de Investigación de Alto Nivel en Odontología, Red Estatal de Odontología num. #REO19–012 (to C.R.).

## Author contribution statement

R.H., M.M., E.N. and C.R. contributed to data acquisition and critically revised the manuscript; A.P., F.M.N. and L.S.S. contributed to MALDI-TOF MS/ML analyses and interpretation, R.H. and C.R. contributed to nLC MS/MS analyses and interpretation. P.A.C. and J.G. contributed to western blotting and vitamin determinations. C.R. contributed to conception, design, project supervision, performed all statistical analyses, and interpretation, drafted and critically revised the manuscript. All authors gave their final approval and agreed to be accountable for all aspects of the work.

## Data availability

All supplementary files are included in this article and are publicly available at: https://doi.org/10.5281/zenodo.4482898. The mass spectrometry proteomic data have been deposited in the ProteomeXchange Consortium via the PRIDE partner repository ^87^ with the dataset identifier PXD026401 (https://www.ebi.ac.uk/pride/archive/projects/PXD026401). All other data supporting the findings of this study are available from the corresponding author on reasonable request.

## REFERENCES

1 Rivera, C. Insights into etiopathogenic and clinical features in 3RU oral diseases: an interesting and challenging research focus. J Oral Res 6, 35 (2017).

2 Edgar, N. R., Saleh, D. & Miller, R. A. Recurrent Aphthous Stomatitis: A Review. J Clin Aesthet Dermatol 10, 26–36 (2017).

3 Jin, L. J. et al. Global burden of oral diseases: emerging concepts, management and interplay with systemic health. Oral Dis 22, 609-619, doi:10.1111/odi.12428 (2016).

4 Lalla, R. V. et al. Multivitamin therapy for recurrent aphthous stomatitis: a randomized, double-masked, placebo-controlled trial. J Am Dent Assoc 143, 370–376 (2012).

5 Rajan, B. et al. Assessment of quality of life in patients with chronic oral mucosal diseases: a questionnaire-based study. Perm J 18, e123-127, doi:10.7812/tpp/13-095 (2014).

6 Akintoye, S. O. & Greenberg, M. S. Recurrent aphthous stomatitis. Dent Clin North Am 58, 281–297, doi:10.1016/j.cden.2013.12.002 (2014).

7 Staines, K. & Greenwood, M. Aphthous ulcers (recurrent). BMJ Clin Evid 2015, 1303 (2015).

8 Swain, S. K., Gupta, S. & Sahu, M. C. Recurrent aphthous ulcers - Still a challenging clinical entity. Apollo Medicine 14, 202–206, doi:10.4103/am.am_40_17 (2017).

9 Eisen, D. & Lynch, D. P. Selecting topical and systemic agents for recurrent aphthous stomatitis. Cutis 68, 201–206 (2001).

10 Rivera, C., Muñoz, A., Puentes, C. & Aguayo, E. Risk Factors for Recurrent Aphthous Stomatitis: A Systematic Review. Preprints, 2021050194, doi:10.20944/preprints202105.0194.v1 (2021).

11 Eguia-del Valle, A., Martinez-Conde-Llamosas, R., Lopez-Vicente, J., Uribarri-Etxebarria, A. & Aguirre-Urizar, J. M. Salivary levels of Tumour Necrosis Factor-alpha in patients with recurrent aphthous stomatitis. Med Oral Patol Oral Cir Bucal 16, e33–36 (2011).

12 Kalpana, R., Thubashini, M. & Sundharam, B. S. Detection of salivary interleukin-2 in recurrent aphthous stomatitis. J Oral Maxillofac Pathol 18, 361–364, doi:10.4103/0973-029x.151313 (2014).

13 Veena, H. R., Mahantesha, S., Joseph, P. A., Patil, S. R. & Patil, S. H. Dissemination of aerosol and splatter during ultrasonic scaling: a pilot study. J Infect Public Health 8, 260–265, doi:10.1016/j.jiph.2014.11.004 (2015).

14 Vucicevic Boras, V. & Savage, N. W. Recurrent aphthous ulcerative disease: presentation and management. Aust Dent J 52, 10-15; quiz 73 (2007).

15 Rivera, C. Essentials of recurrent aphthous stomatitis (Review). Biomed Rep 11, 47–50, doi:10.3892/br.2019.1221 (2019).

16 Moghieb, A., Mangaonkar, M. & Wang, K. K. Mass spectrometry based translational neuroinjury proteomics. Translational Proteomics 1, 65–73 (2013).

17 Belenguer-Guallar, I., Jimenez-Soriano, Y. & Claramunt-Lozano, A. Treatment of recurrent aphthous stomatitis. A literature review. J Clin Exp Dent 6, e168–174, doi:10.4317/jced.51401 (2014).

18 Lamy, E. & Mau, M. Saliva proteomics as an emerging, non-invasive tool to study livestock physiology, nutrition and diseases. J Proteomics 75, 4251–4258, doi:10.1016/j.jprot.2012.05.007 (2012).

19 Belstrom, D. et al. Metaproteomics of saliva identifies human protein markers specific for individuals with periodontitis and dental caries compared to orally healthy controls. PeerJ 4, e2433, doi:10.7717/peerj.2433 (2016).

20 Szabo, G. T. et al. Comparative salivary proteomics of cleft palate patients. Cleft Palate Craniofac J 49, 519–523, doi:10.1597/10-135 (2012).

21 Baldini, C. et al. Proteomic analysis of saliva: a unique tool to distinguish primary Sjogren’s syndrome from secondary Sjogren’s syndrome and other sicca syndromes. Arthritis Res Ther 13, R194, doi:10.1186/ar3523 (2011).

22 Ji, E. H. et al. Potential protein biomarkers for burning mouth syndrome discovered by quantitative proteomics. Mol Pain 13, 1744806916686796, doi:10.1177/1744806916686796 (2017).

23 Winck, F. V. et al. Insights into immune responses in oral cancer through proteomic analysis of saliva and salivary extracellular vesicles. Sci Rep 5, 16305, doi:10.1038/srep16305 (2015).

24 Al-Tarawneh, S. K., Border, M. B., Dibble, C. F. & Bencharit, S. Defining salivary biomarkers using mass spectrometry-based proteomics: a systematic review. Omics 15, 353–361, doi:10.1089/omi.2010.0134 (2011).

25 Williamson, S., Munro, C., Pickler, R., Grap, M. J. & Elswick, R. K., Jr. Comparison of biomarkers in blood and saliva in healthy adults. Nurs Res Pract 2012, 246178, doi:10.1155/2012/246178 (2012).

26 Xiao, H. & Wong, D. T. Method development for proteome stabilization in human saliva. Anal Chim Acta 722, 63–69, doi:10.1016/j.aca.2012.02.017 (2012).

27 Zhu, N. et al. A Novel Coronavirus from Patients with Pneumonia in China, 2019. N Engl J Med 382, 727–733, doi:10.1056/NEJMoa2001017 (2020).

28 Diz, A. P., Truebano, M. & Skibinski, D. O. F. The consequences of sample pooling in proteomics: an empirical study. Electrophoresis 30, 2967–2975, doi:10.1002/elps.200900210 (2009).

29 Meier, F. et al. Online Parallel Accumulation-Serial Fragmentation (PASEF) with a Novel Trapped Ion Mobility Mass Spectrometer. Mol Cell Proteomics 17, 2534–2545, doi:10.1074/mcp.TIR118.000900 (2018).

30 Yu, F. et al. Fast Quantitative Analysis of timsTOF PASEF Data with MSFragger and IonQuant. Mol Cell Proteomics 19, 1575–1585, doi:10.1074/mcp.TIR120.002048 (2020).

31 Choi, M. et al. MSstats: an R package for statistical analysis of quantitative mass spectrometry-based proteomic experiments. Bioinformatics 30, 2524–2526, doi:10.1093/bioinformatics/btu305 (2014).

32 Choi, M. & Tsai, T.-H. Protein significance analysis of mass spectrometry-based proteomics experiments with R and MSstats (v3. 18.1) or later. Msstats <http://msstats.org/wp-content/uploads/2020/02/MSstats_v3.18.1_manual_2020Feb26-v2.pdf/>, [accessed 07 November 2020] (2020).

33 Reimand, J. & Isserlin, R. Pathway enrichment analysis and visualization of omics data using g:Profiler, GSEA, Cytoscape and EnrichmentMap. 14, 482–517, doi:10.1038/s41596-018-0103-9 (2019).

34 Raudvere, U. et al. g:Profiler: a web server for functional enrichment analysis and conversions of gene lists (2019 update). Nucleic Acids Res 47, W191–w198, doi:10.1093/nar/gkz369 (2019).

35 Liao, Y., Wang, J., Jaehnig, E. J., Shi, Z. & Zhang, B. WebGestalt 2019: gene set analysis toolkit with revamped UIs and APIs. Nucleic Acids Res 47, W199–w205, doi:10.1093/nar/gkz401 (2019).

36 Binns, D. et al. QuickGO: a web-based tool for Gene Ontology searching. Bioinformatics 25, 3045–3046, doi:10.1093/bioinformatics/btp536 (2009).

37 Nachtigall, F. M., Pereira, A., Trofymchuk, O. S. & Santos, L. S. Detection of SARS-CoV-2 in nasal swabs using MALDI-MS. 38, 1168–1173, doi:10.1038/s41587-020-0644-7 (2020).

38 Hall, M. A. Correlation-Based Feature Selection for Machine Learning. Ph.D. Thesis University of Waikato, Hamilton, New Zealand (1999).

39 Hall, M. et al. The WEKA data mining software: an update. SIGKDD Explor. Newsl. 11, 10–18, doi:10.1145/1656274.1656278 (2009).

40 Lê, S., Josse, J. & Husson, F. FactoMineR: An R Package for Multivariate Analysis. J Stat Softw 25, 18, doi:10.18637/jss.v025.i01 (2008).

41 Kassambara, A. & Mundt, F. factoextra: Extract and visualize the results of multivariate data analyses. R package version 1.0.3 <https://CRAN.Rproject.org/package=factoextra/,>, [accessed 13 Dec 2020] (2016).

42 Cairns, D. A. et al. Sample size determination in clinical proteomic profiling experiments using mass spectrometry for class comparison. Proteomics 9, 74–86, doi:10.1002/pmic.200800417 (2009).

43 Zhou, C. et al. Statistical considerations of optimal study design for human plasma proteomics and biomarker discovery. J Proteome Res 11, 2103–2113, doi:10.1021/pr200636x (2012).

44 Al-Omiri, M. K. et al. Recurrent aphthous stomatitis (RAS): a preliminary within-subject study of quality of life, oral health impacts and personality profiles. J Oral Pathol Med 44, 278–283, doi:10.1111/jop.12232 (2015).

45 Jurge, S., Kuffer, R., Scully, C. & Porter, S. R. Mucosal disease series. Number VI. Recurrent aphthous stomatitis. Oral Dis 12, 1–21, doi:10.1111/j.1601-0825.2005.01143.x (2006).

46 Dewhirst, F. E. et al. The human oral microbiome. J Bacteriol 192, 5002–5017, doi:10.1128/jb.00542-10 (2010).

47 Chen, T. et al. The Human Oral Microbiome Database: a web accessible resource for investigating oral microbe taxonomic and genomic information. Database (Oxford) 2010, baq013, doi:10.1093/database/baq013 (2010).

48 Scully, C., Gorsky, M. & Lozada-Nur, F. The diagnosis and management of recurrent aphthous stomatitis: a consensus approach. J Am Dent Assoc 134, 200–207, doi:10.14219/jada.archive.2003.0134 (2003).

49 Scully, C. & Porter, S. Oral mucosal disease: recurrent aphthous stomatitis. Br J Oral Maxillofac Surg 46, 198–206, doi:10.1016/j.bjoms.2007.07.201 (2008).

50 Kammers, K., Cole, R. N., Tiengwe, C. & Ruczinski, I. Detecting Significant Changes in Protein Abundance. EuPA Open Proteom 7, 11–19, doi:10.1016/j.euprot.2015.02.002 (2015).

51 Keshamouni, V. G. et al. Differential protein expression profiling by iTRAQ-2DLC-MS/MS of lung cancer cells undergoing epithelial-mesenchymal transition reveals a migratory/invasive phenotype. J Proteome Res 5, 1143–1154, doi:10.1021/pr050455t (2006).

52 Larochelle, M. R. “Is It Safe for Me to Go to Work?” Risk Stratification for Workers during the Covid-19 Pandemic. N Engl J Med, doi:10.1056/NEJMp2013413 (2020).

53 Nalbantoğlu, B. & Nalbantoğlu, A. Vitamin D Levels in Children With Recurrent Aphthous Stomatitis. Ear Nose Throat J 99, 460–463, doi:10.1177/0145561319882783 (2020).

54 Al-Amad, S. H. & Hasan, H. Vitamin D and hematinic deficiencies in patients with recurrent aphthous stomatitis. Clin Oral Investig 24, 2427–2432, doi:10.1007/s00784-019-03102-9 (2020).

55 Al-Maweri, S. A. et al. Is vitamin D deficiency a risk factor for recurrent aphthous stomatitis? A systematic review and meta-analysis. Oral Dis, doi:10.1111/odi.13189 (2019).

56 Subramaniam, P. & Kumar, K. Oral mucosal status and salivary IgA levels of HIV-infected children. J Oral Pathol Med 42, 705–710, doi:10.1111/jop.12061 (2013).

57 Piskin, S., Sayan, C., Durukan, N. & Senol, M. Serum iron, ferritin, folic acid, and vitamin B12 levels in recurrent aphthous stomatitis. J Eur Acad Dermatol Venereol 16, 66–67, doi:10.1046/j.1468-3083.2002.00369.x (2002).

58 Volkov, I., Rudoy, I., Abu-Rabia, U., Masalha, T. & Masalha, R. Case report: Recurrent aphthous stomatitis responds to vitamin B12 treatment. Can Fam Physician 51, 844–845 (2005).

59 Khayamzadeh, M., Najafi, S., Sadrolodabaei, P., Vakili, F. & Kharrazi Fard, M. J. Determining salivary and serum levels of iron, zinc and vitamin B(12) in patients with geographic tongue. J Dent Res Dent Clin Dent Prospects 13, 221–226, doi:10.15171/joddd.2019.034 (2019).

60 Chiang, C. P. et al. Recurrent aphthous stomatitis - Etiology, serum autoantibodies, anemia, hematinic deficiencies, and management. J Formos Med Assoc 118, 1279–1289, doi:10.1016/j.jfma.2018.10.023 (2019).

61 Sun, A. et al. Significant association of deficiencies of hemoglobin, iron, vitamin B12, and folic acid and high homocysteine level with recurrent aphthous stomatitis. J Oral Pathol Med 44, 300–305, doi:10.1111/jop.12241 (2015).

62 Hoffmann, P. R. & Berry, M. J. The influence of selenium on immune responses. Mol Nutr Food Res 52, 1273–1280, doi:10.1002/mnfr.200700330 (2008).

63 Klasser, G. D., Epstein, J. B., Villines, D. & Utsman, R. Burning mouth syndrome: a challenge for dental practitioners and patients. Gen Dent 59, 210-220; quiz 221-212 (2011).

64 Yamamoto, Y. et al. Spinorphin as an endogenous inhibitor of enkephalin-degrading enzymes: roles in pain and inflammation. Curr Protein Pept Sci 3, 587–599, doi:10.2174/1389203023380404 (2002).

65 Kamysz, E., Sałaga, M., Sobocińska, M., Giełdoń, A. & Fichna, J. Anti-inflammatory effect of novel analogs of natural enkephalinase inhibitors in a mouse model of experimental colitis. Future Med Chem 8, 2231–2243, doi:10.4155/fmc-2016-0156 (2016).

66 Gao, S., Zhu, H., Zuo, X. & Luo, H. Cathepsin G and Its Role in Inflammation and Autoimmune Diseases. Arch Rheumatol 33, 498–504, doi:10.5606/ArchRheumatol.2018.6595 (2018).

67 Hoffmann, D. C. et al. Pivotal role for alpha1-antichymotrypsin in skin repair. J Biol Chem 286, 28889–28901, doi:10.1074/jbc.M111.249979 (2011).

68 Bao, J. et al. Combined signatures of serum proteome and transcriptome in patients with recurrent aphthous ulcer. Oral Dis, doi:10.1111/odi.13800 (2021).

69 Sanrattana, W., Maas, C. & de Maat, S. SERPINs-From Trap to Treatment. Front Med (Lausanne) 6, 25, doi:10.3389/fmed.2019.00025 (2019).

70 Yuan, L. et al. Carbonic Anhydrase 1-Mediated Calcification Is Associated With Atherosclerosis, and Methazolamide Alleviates Its Pathogenesis. Front Pharmacol 10, 766, doi:10.3389/fphar.2019.00766 (2019).

71 Henry, E. K. et al. Carbonic anhydrase enzymes regulate mast cell-mediated inflammation. J Exp Med 213, 1663–1673, doi:10.1084/jem.20151739 (2016).

72 Slebioda, Z., Szponar, E. & Kowalska, A. Etiopathogenesis of recurrent aphthous stomatitis and the role of immunologic aspects: literature review. Arch Immunol Ther Exp (Warsz) 62, 205–215, doi:10.1007/s00005-013-0261-y (2014).

73 Natah, S. S., Häyrinen-Immonen, R., Hietanen, J., Malmström, M. & Konttinen, Y. T. Quantitative assessment of mast cells in recurrent aphthous ulcers (RAU). J Oral Pathol Med 27, 124–129, doi:10.1111/j.1600-0714.1998.tb01927.x (1998).

74 Spaans, S. K., Weusthuis, R. A., van der Oost, J. & Kengen, S. W. NADPH-generating systems in bacteria and archaea. Front Microbiol 6, 742, doi:10.3389/fmicb.2015.00742 (2015).

75 Shen, C.-H. in Diagnostic Molecular Biology (ed Chang-Hui Shen) 87–116 (Academic Press, 2019).

76 Opperman, T. & Richardson, J. P. Phylogenetic analysis of sequences from diverse bacteria with homology to the Escherichia coli rho gene. J Bacteriol 176, 5033–5043, doi:10.1128/jb.176.16.5033-5043.1994 (1994).

77 MacNeil, J. R., Blain, A. E., Wang, X. & Cohn, A. C. Current Epidemiology and Trends in Meningococcal Disease-United States, 1996-2015. Clin Infect Dis 66, 1276–1281, doi:10.1093/cid/cix993 (2018).

78 Retchless, A. C. et al. Oropharyngeal microbiome of a college population following a meningococcal disease outbreak. Sci Rep 10, 632, doi:10.1038/s41598-020-57450-8 (2020).

79 Ship, J. A., Chavez, E. M., Doerr, P. A., Henson, B. S. & Sarmadi, M. Recurrent aphthous stomatitis. Quintessence Int 31, 95–112 (2000).

80 Riera Matute, G. & Riera Alonso, E. [Recurrent aphthous stomatitis in Rheumatology]. Reumatol Clin 7, 323–328, doi:10.1016/j.reuma.2011.05.003 (2011).

81 Bankvall, M. et al. The oral microbiota of patients with recurrent aphthous stomatitis. J Oral Microbiol 6, 25739, doi:10.3402/jom.v6.25739 (2014).

82 Stehlikova, Z. et al. Oral Microbiota Composition and Antimicrobial Antibody Response in Patients with Recurrent Aphthous Stomatitis. Microorganisms 7, doi:10.3390/microorganisms7120636 (2019).

83 Kim, Y. J. et al. Mucosal and salivary microbiota associated with recurrent aphthous stomatitis. BMC Microbiol 16 Suppl 1, 57, doi:10.1186/s12866-016-0673-z (2016).

84 Yang, Z. et al. Comparison of microbiomes in ulcerative and normal mucosa of recurrent aphthous stomatitis (RAS)-affected patients. BMC Oral Health 20, 128, doi:10.1186/s12903-020-01115-5 (2020).

85 Hijazi, K. et al. Mucosal microbiome in patients with recurrent aphthous stomatitis. J Dent Res 94, 87s–94s, doi:10.1177/0022034514565458 (2015).

86 Grassl, N. et al. Ultra-deep and quantitative saliva proteome reveals dynamics of the oral microbiome. Genome Med 8, 44, doi:10.1186/s13073-016-0293-0 (2016).

87 Perez-Riverol, Y. et al. The PRIDE database and related tools and resources in 2019: improving support for quantification data. Nucleic Acids Res 47, D442–d450, doi:10.1093/nar/gky1106 (2019).

