## Supplementary file for "Salivary proteome of aphthous stomatitis reveals the participation of vitamin metabolism, nutrients, and bacteria"

**SUPPLEMENTARY MATERIAL**

**Supplementary methods**

- Quality of salivary samples.
- Protein extraction and trypsin digestion.
- Liquid chromatography-tandem mass spectrometry.
- Mass spectrometry data analysis.
- MALDI-MS.

**Supplementary tables**

- Supplementary Table S1. Clinical characteristics of participants.
- Supplementary Table S2. List of diseases and conditions for exclusion.
- Supplementary Table S3. Showing detailed pools.
- Supplementary Table S4. Peaks from proteomic profiles selected with CFS for PCA graphics in in Figure 5.

**Supplementary figure**

- Supplementary Figure S1. Uncropped pictures of gels and blots.

**Supplementary methods**

**Quality of salivary samples.** To evaluate the quality of the samples, we prepared 2 polyacrylamide gels of 10 lanes each with a thickness of 1.5 mm. Then, we added 20 ug protein to each lane, with loading buffer (Pierce™ Lane Marker Reducing Sample Buffer, # 1610373, Thermo Scientific). As internal control we used 20 ug of depleted human serum (Pierce™ Top 2 Abundant Protein Depletion Spin Columns, # 85161 Thermo Fisher Scientific). The samples were subjected to electrophoresis under denaturing conditions (SDS-PAGE) on a 10% gel, where they were run at 100V and at the end at 120V on ice. Gel was stained overnight with 50 mL of Coomassie blue R-250 (# 1610786, Bio-Rad) while shaking at room temperature. The photo documentation of the gels was run on an Odyssey® CLx Imaging System equipment.

**Protein extraction and trypsin digestion.** Proteins were precipitated by adding 5 volumes of cold acetone and incubated overnight at -80 °C. Then, they were equilibrated at room temperature for 10 min, they were centrifuged at 16,000 x g for 15 min at 4 ° C and the supernatant was discarded. The resulting pellet was washed 3 times with cold 80% acetone. Subsequently, the protein pellet was allowed to dry in a rotary concentrator.

Samples were resuspended in 30 μL 8M Urea and 25 mM ammonium bicarbonate. Then, they were reduced with DTT to a final concentration of 20 mM in 25 mM ammonium bicarbonate and incubated for 1 hour at room temperature. Then, they were alkylated by adding iodoacetamide to a final concentration of 20 mM in 25 mM ammonium bicarbonate and incubated for 1 hour in the dark at room temperature. Subsequently, the samples were diluted 8 times with 25 mM ammonium bicarbonate.

Digestion was performed with sequencing grade trypsin (#V5071, Promega) in a 1:50 ratio of protease: protein (mass/mass), incubated for 16 h at 37 ° C. Digestion reaction was stopped by pH adding formic acid to the 10%. The samples were then subjected to Clean Up SepPack Vac C-18 (Waters), according to the manufacturer's instructions. Subsequently, clean peptides were dried in a rotary concentrator at 1,000 rpm overnight at 10°C. The pure and dry peptides were resuspended in 10 μL of 0.1% formic acid and then quantified in Direct Detect IR (Millipore), leaving a dilution of 0.5 μg/μl, for injection in the timsTOF Pro mass spectrometer (Bruker Daltonics).

**Liquid chromatography-tandem mass spectrometry.** A complex mixture of tryptic peptide derived from salivary samples was diluted with 0.1% formic acid to 0.5 µg/µL concentration. We injected 500 ng of peptides into a nanoElute liquid chromatography system (Bruker Daltonics) coupled to a timsTOF Pro mass spectrometer (Bruker Daltonics) using an Aurora UHPLC column (25cm x 75μm ID, 1.6μm C18, IonOpticks). Chromatographic separation was performed using a 90 minutes linear gradient of 2% to 35% of buffer B (0.1% formic acid in acetonitrile). Next, we collected MS data using the oTOF software (Bruker Daltonics) under 10 PASEF cycles with an m/z range of 100 to 1,700. The capillary ionization was 1,600 V and its temperature was 200°C. TOF frequency was 10 KHz at a resolution of 50,000 FWHM.

**Mass spectrometry data analysis.** Mass spectrometry raw files were processed with MsFragger ^1^. For all searches, a protein sequence database of reviewed human proteins (accessed 06/15/2020 from UniProt; 74,823 entries) was used. Decoy sequences were generated and appended to the original database using Philosopher. We did an open and semi-tryptic search. A maximum of two missing cleavages were allowed, the required minimum peptide sequence length was 7 amino acids, and the peptide mass was limited to a maximum of 5,000 Da. Carbamidomethylation of cysteine residues was set as a fixed modification. Methionine oxidation (M), acetylation of protein N termini ([^), pyro-glutamic acid and pyro-carbamidomethyl cysteine (nQnC) and water loss on any peptide N-terminus (NE) as variable modifications. Precursor mass tolerance was set from -150 to +500 Da, and precursor true tolerance and fragment mass tolerance were set to 20 ppm. Mass calibration and parameter optimization were enabled. Two missed cleavages were allowed, and the number of enzymatic termini was set to two. Isotope error was set to 0. The minimum number of fragment peaks required to include a PSM in modelling was set to two, and the minimum number required reporting the match was four. A minimum of 15 fragment peaks and the top 100 most intense peaks were used as initial settings. In IonQuant, we selected match between runs (MBR), mass tolerance was set to 10 ppm, retention time tolerance was set to 0.4 minutes, ion mobility (1/K0) tolerance was set to 0.05, normalization was enabled, and minimum isotope count was set to 2 by default. Minimum ion counts 2 was selected.

**MALDI-MS/ML.** Mass spectrometric analyses were performed with a MALDI time-of-flight instrument (Autoflex, Bruker) with a pulsed nitrogen laser (337 nm), operating in positive-ion linear mode using a 19 kV acceleration voltage. The matrix solution was prepared with α-ciano-hydroxy-cinnamic acid (CHCA) at 1% in acetonitrile/0.1% trifluoroacetic acid (1:1). One microliter of each saliva swab sample was spotted on a MALDI steel plate followed by the addition of 1 μl of the matrix solution (CHCA) and air-drying. Spectra were generated by summing 500 single spectra (10 × 50 shots) in the range between 3 and 20 kDa by shooting the laser at random positions on the target spot. MALDI-MS fid files (Bruker) were converted to mzML with MSconvert from the ProteoWizard suit ^2^, and subsequently preprocessed in R environment using the MALDIquant and MALDIquantForeign packages ^3^. All spectra were trimmed to a range from 3 to 18 kDa. Square root transformation was applied, and smoothing was realized by the Savitzky–Golay method. The baseline correction was performed using the SNIP algorithm, and the intensity was normalized using the total ion current calibration method. Spectra were aligned with a tolerance of 0.002. Peak detection was carried out applying a signal-to-noise ratio of 4 and a halfWindowSize of 20. Peaks were binned through the binpeaks command with a tolerance of 0.002, and then the matrix of peak intensities was generated.

The most relevant peaks were selected with the correlation-based feature subset selection (CFS) method ^4^ implemented in Weka software ^5^. To explore and compare spectra in multidimensional space, principal component analysis (PCA) was conducted using the R FactoMineR ^6^ and factoextra ^7^ packages (data were scaled to unit variance). The following comparisons were made: ulcerative stage vs healthy contralateral, remission stage healed ulcer vs healthy contralateral, recurrence of ulcer vs healthy contralateral and healthy controls vs healthy contralateral ulcerative stage vs remission stage healed ulcer vs remission stage healthy contralateral vs recurrence of ulcer healthy contralateral. In the last one, the selection of peaks by CFS was not performed because only 1 attribute was retained, so for this PCA all the peaks were used.

**Supplementary tables**

**Supplementary Table S1.** Clinical characteristics of participants.

| **Ulcerative stage** | | | | | | | | |
| --- | --- | --- | --- | --- | --- | --- | --- | --- |
| **Pat.** | **Sex** | **Age ranges** | **Epi. year** | **Ulcers num.** | **Size (mean)** | **Ove. dia** | **OHIP** | **Location** |
| A1 | H | 20-29 | 4 | 2 | 1.5 | 3 | 8 | C06.1 Vestibule of mouth |
| A2 | M | ≤19 | 0 | 1 | 2 | 2 | 3 | C06.1 Vestibule of mouth |
| A3 | M | ≤19 | 4 | 1 | 4 | 4 | 17 | C06.0 Cheek mucosa |
| A4 | M | 20-29 | 12 | 1 | 3 | 3 | 19 | C00.4 Mucosa of lower lip |
| A5 | M | ≥40 | 1 | 1 | 5 | 5 | 21 | C06.1 Vestibule of mouth |
| A6 | M | 20-29 | 3 | 1 | 2 | 2 | 33 | C02.1 Border of tongue |
| A7 | M | 20-29 | 20 | 2 | 3 | 4 | 21 | C02.0 Dorsal surface of tongue, NOS + C06.0 Cheek mucosa |
| A8 | M | 20-29 | 0 | 2 | 2 | 3 | 13 | C00.4 Mucosa of lower lip |
| A9 | M | 20-29 | 0 | 1 | 3 | 3 | 3 | C00.4 Mucosa of lower lip |
| A10 | M | 20-29 | 5 | 1 | 3 | 3 | 19 | C02.1 Border of tongue |
| A11 | M | ≥40 | 6 | 1 | 4 | 4 | 26 | C00.4 Mucosa of lower lip |
| A12 | M | 20-29 | 2 | 1 | 2 | 2 | 24 | C00.4 Mucosa of lower lip |
| A13 | H | 30-39 | 2 | 2 | 1 | 2 | 5 | C06.1 Vestibule of mouth |
| A14 | H | 30-39 | 2 | 2 | 2 | 3 | 7 | C06.1 Vestibule of mouth |
| A15 | M | ≤19 | 8 | 1 | 3 | 3 | 8 | C00.4 Mucosa of lower lip |
| A16 | M | 20-29 | 2 | 1 | 3 | 3 | 38 | C00.4 Mucosa of lower lip |
| A17 | M | 20-29 | 8 | 2 | 5 | 9 | 29 | C04.9 Floor of mouth, NOS + C06.0 Cheek mucosa |
| A18 | M | 20-29 | 12 | 1 | 6 | 6 | 19 | C04.9 Floor of mouth, NOS |
| A19 | M | 20-29 | 6 | 1 | 5 | 5 | 30 | C00.4 Mucosa of lower lip |
| A20 | M | 20-29 | 3 | 1 | 1 | 1 | 6 | C06.0 Cheek mucosa |
| A21 | M | 20-29 | 0 | 1 | 4 | 4 | 14 | C06.1 Vestibule of mouth |
| A22 | M | ≤19 | 10 | 1 | 4 | 4 | 24 | C02.2 Ventral surface of tongue, NOS |
| A23 | M | 20-29 | 5 | 1 | 7 | 7 | 22 | C00.4 Mucosa of lower lip |
| A24 | M | 20-29 | 2 | 1 | 2 | 2 | 16 | C06.0 Cheek mucosa |
| A25 | M | 20-29 | 10 | 1 | 3 | 3 | 11 | C02.2 Ventral surface of tongue, NOS |
| A26 | M | ≥40 | 6 | 3 | 3.3 | 10 | 36 | C00.3 Mucosa of upper lip |
| A27 | M | 20-29 | 0 | 1 | 5 | 5 | 9 | C05.1 Soft palate, NOS |
| A28 | H | 20-29 | 8 | 2 | 2.5 | 5 | 20 | C00.3 Mucosa of upper lip + C00.4 Mucosa of lower lip |
| A29 | M | 20-29 | 24 | 2 | 3 | 3 | 14 | C03.1 Lower gum |
| A30 | M | 20-29 | 0 | 1 | 2 | 2 | 22 | C03.0 Upper gum |
| A31 | H | 20-29 | 15 | 1 | 3 | 3 | 20 | C05.1 Soft palate, NOS |
| A32 | M | ≤19 | 2 | 1 | 3 | 3 | 25 | C06.0 Cheek mucosa |
| A33 | H | ≥40 | 12 | 4 | 5 | 5 | 21 | C06.1 Vestibule of mouth |
| A34 | M | ≥40 | 12 | 2 | 3 | 6 | 34 | C00.4 Mucosa of lower lip + C06.1 Vestibule of mouth |
| A35 | M | 20-29 | 3 | 1 | 1 | 1 | 9 | C06.1 Vestibule of mouth |
| A36 | M | 20-29 | 0 | 3 | 0.5 | 1.5 | 12 | C03.1 Lower gum |
| A37 | M | 20-29 | 0 | 2 | 2 | 4 | 11 | C03.1 Lower gum |
| **Recurrence** | | | | | | | | |
| **Pat.** | **Sex** | **Age ranges** | **Epi. year** | **Ulcers num.** | **Size (mean)** | **Ove. dia** | **OHIP** | **Location** |
| R1 | M | 20-29 | 20 | 1 | 1 | 1 | 9 | - |
| R2 | M | ≥40 | 1 | 1 | 1 | 1 | 13 | C06.1 Vestibule of mouth |
| R3 | M | 20-29 | 12 | 1 | 3 | 3 | 17 | C03.1 Lower gum |
| R4 | M | 20-29 | 6 | 2 | 5 | 8 | 23 | C00.4 Mucosa of lower lip |
| R5 | M | 20-29 | 2 | 1 | 3 | 3 | 4 | C00.4 Mucosa of lower lip |
| R6 | H | 20-29 | 4 | 1 | 3 | 3 | 5 | C00.4 Mucosa of lower lip |
| R7 | M | ≤19 | 4 | 1 | 1 | 1 | 1 | C00.4 Mucosa of lower lip |
| R8 | M | ≥40 | 6 | 2 | 3 | 5 | 17 | C00.4 Mucosa of lower lip |
| R9 | M | 20-29 | 12 | 1 | 5 | 5 | 25 | C00.4 Mucosa of lower lip |
| R10 | M | 20-29 | 3 | 1 | 1 | 1 | 6 | C00.4 Mucosa of lower lip |
| R11 | M | 20-29 | 2 | 1 | 1 | 1.5 | 15 | C00.3 Mucosa of upper lip |
| R12 | M | 20-29 | 3 | 1 | 1 | 1 | 6 | C00.4 Mucosa of lower lip |
| R13 | M | 20-29 | 20 | 1 | 2 | 2 | 9 | C06.0 Cheek mucosa |
| R14 | H | 30-39 | 2 | 1 | 2 | 2 | 4 | C05.1 Soft palate, NOS |
| R15 | H | 30-39 | 2 | 1 | 2 | 2 | 3 | C05.1 Soft palate, NOS |
| **Remission stage** | | | | | | | | |
| **Pat.** | **Sex** | **Age ranges** | **Epi. year** | **Ulcers num.** | **Size (mean)** | **Ove. dia** | **OHIP** | **Location** |
| D1 | M | ≤19 | 4 | 0 | 0 | 0 | 6 | - |
| D2 | H | 20-29 | 4 | 0 | 0 | 0 | 5 | - |
| D3 | M | 20-29 | 12 | 0 | 0 | 0 | 3 | - |
| D4 | M | 20-29 | 0 | 0 | 0 | 0 | 3 | - |
| D5 | M | ≤19 | 0 | 0 | 0 | 0 | 4 | - |
| D6 | M | 20-29 | 0 | 0 | 0 | 0 | 7 | - |
| D7 | M | ≥40 | 6 | 0 | 0 | 0 | 15 | - |
| D8 | M | 20-29 | 20 | 0 | 0 | 0 | 8 | - |
| D9 | M | 20-29 | 6 | 0 | 0 | 0 | 5 | - |
| D10 | M | 20-29 | 3 | 0 | 0 | 0 | 16 | - |
| D11 | H | 30-39 | 2 | 0 | 0 | 0 | 1 | - |
| D12 | M | ≤19 | 2 | 0 | 0 | 0 | 10 | - |
| D13 | M | ≤19 | 8 | 0 | 0 | 0 | 7 | - |
| D14 | M | 20-29 | 0 | 0 | 0 | 0 | 8 | - |
| D15 | M | 20-29 | 2 | 0 | 0 | 0 | 11 | - |
| D16 | M | 20-29 | 3 | 0 | 0 | 0 | 9 | - |
| D17 | M | 20-29 | 8 | 0 | 0 | 0 | 21 | - |
| D18 | M | 20-29 | 5 | 0 | 0 | 0 | 1 | - |
| D19 | M | 20-29 | 2 | 0 | 0 | 0 | 2 | - |
| D20 | M | 20-29 | 6 | 0 | 0 | 0 | 14 | - |
| D21 | M | 20-29 | 3 | 0 | 0 | 0 | 2 | - |
| D22 | M | 20-29 | 3 | 0 | 0 | 0 | 11 | - |
| D23 | M | 20-29 | 3 | 0 | 0 | 0 | 8 | - |
| D24 | M | ≤19 | 10 | 0 | 0 | 0 | 10 | - |
| D25 | M | 20-29 | 24 | 0 | 0 | 0 | 3 | - |
| D26 | H | 20-29 | 15 | 0 | 0 | 0 | 10 | - |
| D27 | H | 20-29 | 8 | 0 | 0 | 0 | 3 | - |
| D28 | M | 20-29 | 12 | 0 | 0 | 0 | 12 | - |
| D29 | M | ≥40 | 1 | 0 | 0 | 0 | 4 | - |
| D30 | M | ≥40 | 6 | 0 | 0 | 0 | 9 | - |
| D31 | M | 20-29 | 10 | 0 | 0 | 0 | 3 | - |
| D32 | M | ≤19 | 7 | 0 | 0 | 0 | 17 | - |
| D33 | M | 20-29 | 8 | 0 | 0 | 0 | 9 | - |
| D34 | M | 20-29 | 0 | 0 | 0 | 0 | 7 | - |
| D35 | M | 20-29 | 0 | 0 | 0 | 0 | 8 | - |
| D36 | H | 30-39 | 2 | 0 | 0 | 0 | 1 | - |
| **Healthy controls** | | | | | | | | |
| **Pat.** | **Sex** | **Age ranges** | **Epi. year** | **Ulcers num.** | **Size (mean)** | **Ove. dia** | **OHIP** | **Location** |
| C1 | M | 20-29 | 0 | 0 | 0 | 0 | 17 | - |
| C2 | M | 20-29 | 0 | 0 | 0 | 0 | 14 | - |
| C3 | M | ≤19 | 0 | 0 | 0 | 0 | 13 | - |
| C4 | M | 20-29 | 0 | 0 | 0 | 0 | 17 | - |
| C5 | M | 20-29 | 0 | 0 | 0 | 0 | 9 | - |
| C6 | M | 20-29 | 0 | 0 | 0 | 0 | 11 | - |
| C7 | H | 20-29 | 0 | 0 | 0 | 0 | 14 | - |
| C8 | M | 20-29 | 0 | 0 | 0 | 0 | 12 | - |
| C9 | M | 20-29 | 0 | 0 | 0 | 0 | 7 | - |
| C10 | H | 20-29 | 0 | 0 | 0 | 0 | 5 | - |
| C11 | M | 30-39 | 0 | 0 | 0 | 0 | 7 | - |
| C12 | M | 20-29 | 0 | 0 | 0 | 0 | 12 | - |
| C13 | M | 20-29 | 0 | 0 | 0 | 0 | 5 | - |
| C14 | M | 20-29 | 0 | 0 | 0 | 0 | 15 | - |
| C15 | M | 20-29 | 0 | 0 | 0 | 0 | 10 | - |
| C16 | M | 20-29 | 0 | 0 | 0 | 0 | 10 | - |
| C17 | H | 30-39 | 0 | 0 | 0 | 0 | 12 | - |
| C18 | M | 20-29 | 0 | 0 | 0 | 0 | 7 | - |
| C19 | M | 20-29 | 0 | 0 | 0 | 0 | 23 | - |
| C20 | M | 20-29 | 0 | 0 | 0 | 0 | 17 | - |
| C21 | M | ≤19 | 0 | 0 | 0 | 0 | 11 | - |
| C22 | M | 20-29 | 0 | 0 | 0 | 0 | 4 | - |
| C23 | M | ≥40 | 0 | 0 | 0 | 0 | 18 | - |
| C24 | M | ≥40 | 0 | 0 | 0 | 0 | 5 | - |
| C25 | M | 20-29 | 0 | 0 | 0 | 0 | 20 | - |
| C26 | M | 20-29 | 0 | 0 | 0 | 0 | 4 | - |
| C27 | M | 30-39 | 0 | 0 | 0 | 0 | 5 | - |
| C28 | H | ≤19 | 0 | 0 | 0 | 0 | 11 | - |
| C29 | M | 20-29 | 0 | 0 | 0 | 0 | 9 | - |
| C30 | M | 20-29 | 0 | 0 | 0 | 0 | 3 | - |
| C31 | M | 20-29 | 0 | 0 | 0 | 0 | 11 | - |

Pat. patients; Epi. year, episodes of ulcers in the past year; Num. ulcers, number of ulcers on clinical examination; Ove. dia., (sum of the diameters of ulcerated mucosa); OHIP, OHIP14-SP, impact of oral health on quality of life. The anatomical location was defined according to the codes of the International Classification of Diseases for Oncology (ICD-O-3).

**Supplementary Table S2.** List of diseases and conditions for exclusion.

| **Diseases and conditions** | | |
| --- | --- | --- |
| Behçet syndrome | Oral herpes | Malabsorption |
| Folic acid deficiency | Herpangina | Pernicious anemia |
| Iron deficiency anemia | Lack of iron | Sweet's syndrome (acute febrile neutrophilic dermatosis) |
| Vitamin B12 deficiency | Systemic lupus erythematosus | Marshall syndrome or PFAPA syndrome (periodic fever, aphthous stomatitis, pharyngitis and adenitis) |
| Crohn's disease | Reactive arthritis | Trisomy 8 |
| Ulcerative colitis | Oral candidiasis | Severe periodontal disease |
| Lichen planus | Celiac Disease | Erythema multiforme |
| Pemphigus | Cyclical neutropenia | Syphilis |
| Pemphigoid | Gluten-sensitive enteropathy | HIV-AIDS |

**Supplementary Table S3**. Showing detailed pools.

**Healthy controls**

1: C1, C5, C13, C17, C18, C19, C20, C21, C26, C27, C28

2: C2, C4, C8, C10, C11, C12, C16, C23, C22, C25, C31

3: C2, C3, C5, C6, C7, C9, C14, C15, C24, C29, C30

**Ulcerative stage**

1: A6, A10, A11, A13, A19, A21, A22, A26, A29, A31, A33, A35

2: A1, A2, A9, A12, A14, A15, A20, A25, A27, A28, A30, A36

3: A4, A5, A7, A8, A16, A17, A18, A23, A24, A32, A34, A36

**Remission stage**

1: D7, D8, D9, D12, D13, D16, D17, D21, D25, D26, D29, D34

2: D2, D3, D4, D6, D18, D19, D22, D23, D27, D28, D31, D36

3: D1, D5, D10, D11, D14, D15, D20, D24, D30, D32, D33, D35

**Supplementary Table S4**. Peaks from proteomic profiles selected with CFS for PCA graphics in Figure 5.

| **#** | **Peaks used in Figure 5A** | **Peaks used in Figure 5B** | **Peaks used in Figure 5C** |
| --- | --- | --- | --- |
|  | **Ulcerative stage vs Healthy contralateral** | **Remission stage healed ulcer vs Healthy contralateral** | **Recurrence of ulcer vs Healthy contralateral** |
| 1 | 3234 | 3411 | 3349 |
| 2 | 3349 | 5684 | 3593 |
| 3 | 3392 | 6084 | 3599 |
| 4 | 3486 | 6274 | 3609 |
| 5 | 3528 | 6642 | 3620 |
| 6 | 3573 | 10190 | 3673 |
| 7 | 3614 | 16804 | 3706 |
| 8 | 3620 |  | 4380 |
| 9 | 3680 |  | 5028 |
| 10 | 3748 |  | 8254 |
| 11 | 4117 |  | 11185 |
| 12 | 4386 |  |  |
| 13 | 4554 |  |  |
| 14 | 4591 |  |  |
| 15 | 5357 |  |  |
| 16 | 5913 |  |  |
| 17 | 6252 |  |  |
| 18 | 7325 |  |  |
| 19 | 7588 |  |  |
| 20 | 12904 |  |  |
| 21 | 16010 |  |  |
| * In Figure 5D, peaks selection by CFS was not performed because only 1 attribute was retained, that is why for respective PCA we used all peaks. | | | |

**Supplementary figure**

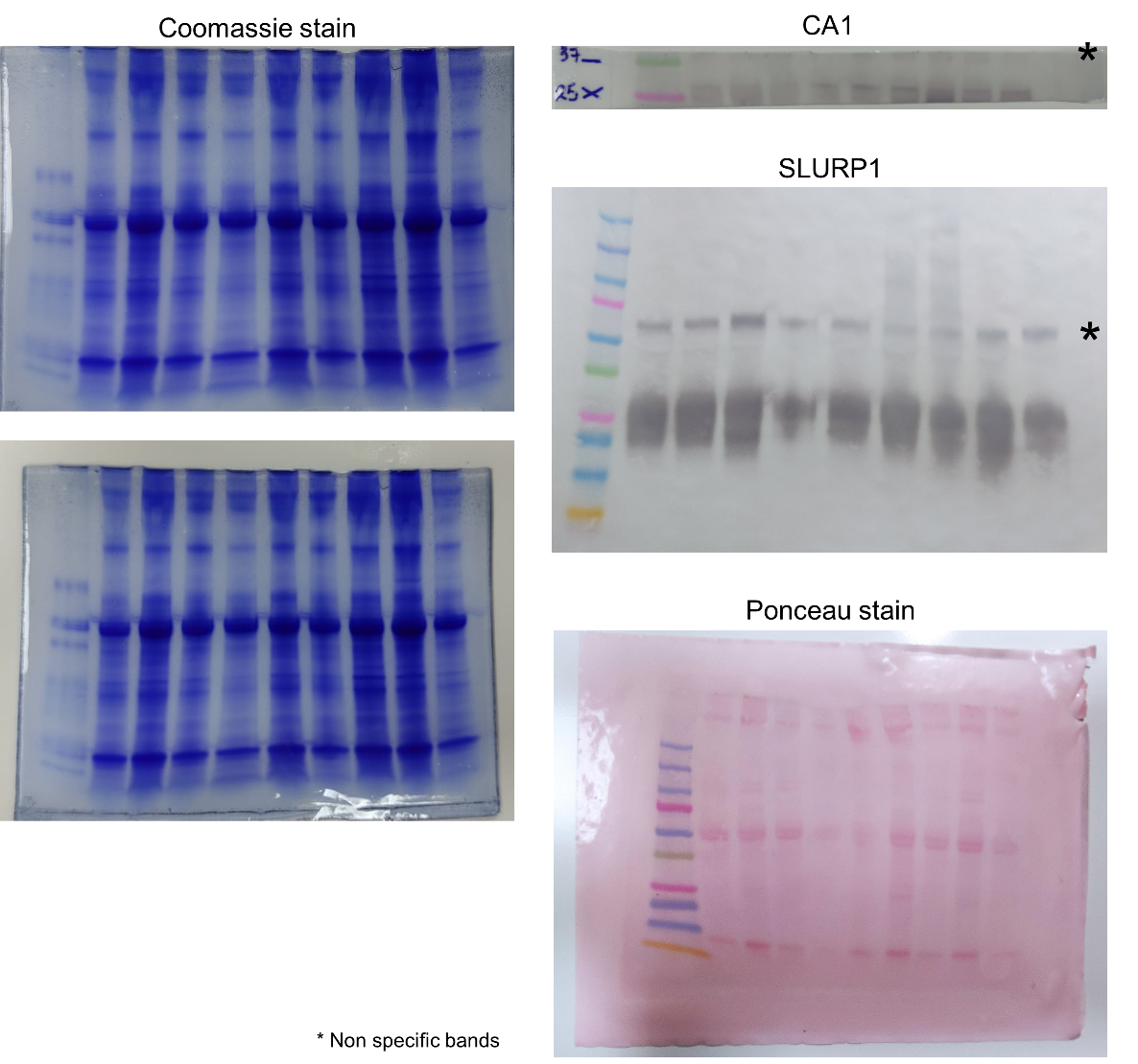

**Supplementary Figure S1.** Uncropped pictures of gels and blots.

**REFERENCES**

1	Yu, F. *et al.* Fast Quantitative Analysis of timsTOF PASEF Data with MSFragger and IonQuant. *Mol Cell Proteomics* **19**, 1575-1585, doi:10.1074/mcp.TIR120.002048 (2020).

2	Chambers, M. C. *et al.* A cross-platform toolkit for mass spectrometry and proteomics. *Nat Biotechnol* **30**, 918-920, doi:10.1038/nbt.2377 (2012).

3	Gibb, S. & Strimmer, K. MALDIquant: a versatile R package for the analysis of mass spectrometry data. *Bioinformatics* **28**, 2270-2271, doi:10.1093/bioinformatics/bts447 (2012).

4	Hall, M. A. Correlation-Based Feature Selection for Machine Learning. *Ph.D. Thesis* **University of Waikato, Hamilton, New Zealand** (1999).

5	Hall, M. *et al.* The WEKA data mining software: an update. *SIGKDD Explor. Newsl.* **11**, 10–18, doi:10.1145/1656274.1656278 (2009).

6	Lê, S., Josse, J. & Husson, F. FactoMineR: An R Package for Multivariate Analysis. *J Stat Softw* **25**, 18, doi:10.18637/jss.v025.i01 (2008).

7	Kassambara, A. & Mundt, F. factoextra: Extract and visualize the results of multivariate data analyses. *R package version 1.0.3* **<**[**https://CRAN.R-project.org/package=factoextra**](https://CRAN.R-project.org/package=factoextra) **/>, [accessed 13 Dec 2020]** (2016).
